# Antimicrobial resistance prevalence in clinical and aquatic environmental ESKAPE: a systematic review with meta-analysis

**DOI:** 10.64898/2026.02.25.26346099

**Authors:** Aline Bruna Martins Vaz, Bruno Murad, Bruna Coelho Lopes, Múcio Leão Pessoa de Castro, Gabriel da Rocha Fernandes, Wanderson Kleber de Oliveira, Paula Luize Camargos Fonseca, Eric Roberto Guimarães Rocha Aguiar, César Rossas Mota Filho, André Bezerra Santos, Carlos Ernesto Ferreira Starling

**Author notes:** Corresponding author: A. B. M. Vaz: Department of Medical Education Development. Universidade Professor Edson Antônio Velano (UNIFENAS) Medical School. Belo Horizonte (Brazil). Full postal address: Líbano street, 66, zip code 31.710-030, Belo Horizonte, Brazil.

## Abstract

Antimicrobial resistance (AMR) in ESKAPE pathogens represents a major global health threat. Although these organisms are well established as causes of healthcare-associated infections, aquatic environments may function as reservoirs and transmission pathways for resistance. This systematic review aimed to estimate the prevalence of AMR in ESKAPE pathogens isolated from water and wastewater and to compare resistance patterns with those observed in human clinical isolates. The review followed PRISMA guidelines and was registered in PROSPERO (CRD420251020930). PubMed, Embase, and the Cochrane Library were searched to January 14, 2025. Eligible studies were original research reporting antimicrobial susceptibility data for ESKAPE pathogens isolated from both aquatic environmental matrices and clinical samples. Pooled resistance prevalence was estimated using generalized linear mixed models, with heterogeneity assessed using τ² and I² statistics and small-study effects evaluated by funnel plots and Egger’s test. Of 304 records identified, 18 studies met the inclusion criteria. The pooled overall resistance prevalence was 0.46 (95% CI: 0.36–0.57), with heterogeneity (I² = 98.8%). Resistance was higher in clinical isolates (0.67; 95% CI: 0.55–0.77) than in environmental isolates (0.24; 95% CI: 0.14–0.39), and environmental resistance was greater in effluent-impacted waters than in non-effluent sources. Interpretation is limited by methodological heterogeneity, selective isolation approaches in environmental studies, and imprecision due to small and unevenly distributed samples. Overall, AMR in ESKAPE pathogens remains more prevalent in clinical settings, but aquatic environments, particularly wastewater, represent resistance reservoirs, underscoring the need for standardized methodologies within a One Health framework.

**Systematic review registration:** https://www.crd.york.ac.uk/PROSPERO/view/CRD420251020930, CRD420251020930

**Highlights:** Antimicrobial resistance was higher in clinical isolates than in aquatic isolates.

Resistance patterns showed extreme heterogeneity across studies.

Effluent-impacted waters showed higher resistance than non-effluent sources.

Higher environmental resistance in some classes reflected methodological artifacts.

## 1. Introduction

Antibiotics are bioactive compounds capable of inhibiting bacterial growth or causing cell death through mechanisms such as disruption of cell wall or membrane integrity, inhibition of protein or nucleic acid synthesis, and selective toxicity toward microbial targets [1]. However, the widespread and often inappropriate use of these agents has intensified selective pressure on microorganisms, enabling them to develop antimicrobial resistance (AMR), defined as the capacity to survive and proliferate despite exposure to drugs that were previously effective [2]. Today, AMR poses a clear and present danger to global public health, threatening to roll back a century of medical advancements [3].

The molecular strategies underlying AMR are both varied and highly effective. Bacteria employ defenses such as producing enzymes to neutralize antibiotics, altering cell walls to limit drug entry, modifying molecular targets or developing alternative metabolic routes [1,2]. Through these strategies, resistant pathogens are able to persist and circulate not only within hospitals but across communities, making infectious diseases increasingly difficult to manage.

Resistance often begins with a spontaneous genetic mutation, which may become dominant within pathogen populations after the introduction of new antimicrobial agents, although this timeline varies across species and drugs [4]. Evolutionary pressure for resistance is amplified in environments where microbes are exposed to low, non-lethal concentrations of antibiotics, a common scenario in agricultural areas, wastewater treatment plants, and natural aquatic systems [3,5]. In clinic settings, this process is further accelerated by inappropriate prescribing, incomplete treatment courses, and the circulation of substandard medicines [2,3]. This interaction among human, animal, and environmental determinants highlights the necessity of a One Health approach to address antimicrobial resistance effectively [6,7].

Recognizing this growing threat, the World Health Organization (WHO) established its Bacterial Priority Pathogens List (BPPL), which ranks pathogens according to their risk to human health [8]. Among these, the ESKAPE pathogens (Enterococcus faecium, Staphylococcus aureus, Klebsiella pneumoniae, Acinetobacter baumannii, Pseudomonas aeruginosa, and Enterobacter spp.) are notable for causing severe healthcare-associated infections and for their capacity to acquire and disseminate resistance genes [9–11]. These organisms pose a particular threat to vulnerable patient populations.

Although ESKAPE pathogens have long been recognized as major clinical challenges, there is increasing evidence that they are widely distributed beyond hospital settings, including in soil [12], food products [13], and aquatic environments such as rivers [14], drinking water sources [15] and sewage systems [13,16]. Aquatic environments, in particular, act as important reservoirs and transmission pathways for AMR, as they continuously receive inputs from wastewater effluent, agricultural runoff, agriculture runoff, and animal waste, facilitating the persistence and spread of resistant bacteria [13,17].

Because human, animal and environmental health are intrinsically interconnected, a One Health framework is essential for a comprehensive understanding of AMR. Within this context, identifying environmental reservoirs and comparing resistance patterns across ecological compartments are critical for understanding transmission pathways [18,19].

Accordingly, this systematic review and meta-analysis aimed to evaluate the prevalence of AMR in ESKAPE pathogens from water and wastewater and to compare these estimates with resistance patterns observed in clinical isolates.

## 2. Methods

This systematic review under the Preferred Reporting Items for Systematic Reviews and Meta-Analyses (PRISMA) guidelines [20,21]. A prospective protocol was formulated and uploaded to PROSPERO (CRD420251020930).

### 2.1 Search strategy and study selection

This systematic review was conducted in accordance with the Preferred Reporting Items for Systematic Reviews and Meta-Analyses (PRISMA) guidelines. A comprehensive literature search was performed in PubMed, Embase, and the Cochrane Library to identify all relevant studies published up to January 14, 2025, selected for their broad coverage of biochemical and environmental health literature.

The search strategy combined keywords and subject headings related to antimicrobial resistance, ESKAPE pathogens, and aquatic environments (Supplementary Table 1). No language or publication date restrictions were applied. All records were imported into Zotero for bibliographic management, and duplicates were removed.

Reference lists of included articles were manually screened to identify additional relevant studies. Study selection was conducted in two phases: initial screening of titles and abstracts by four independent reviewers, followed by full-text assessment by two reviewers.

Disagreements were resolved through discussion and consensus. The complete selection process and reasons for exclusion are presented in the PRISMA flow diagram (Figure 1).

**Figure 1.**
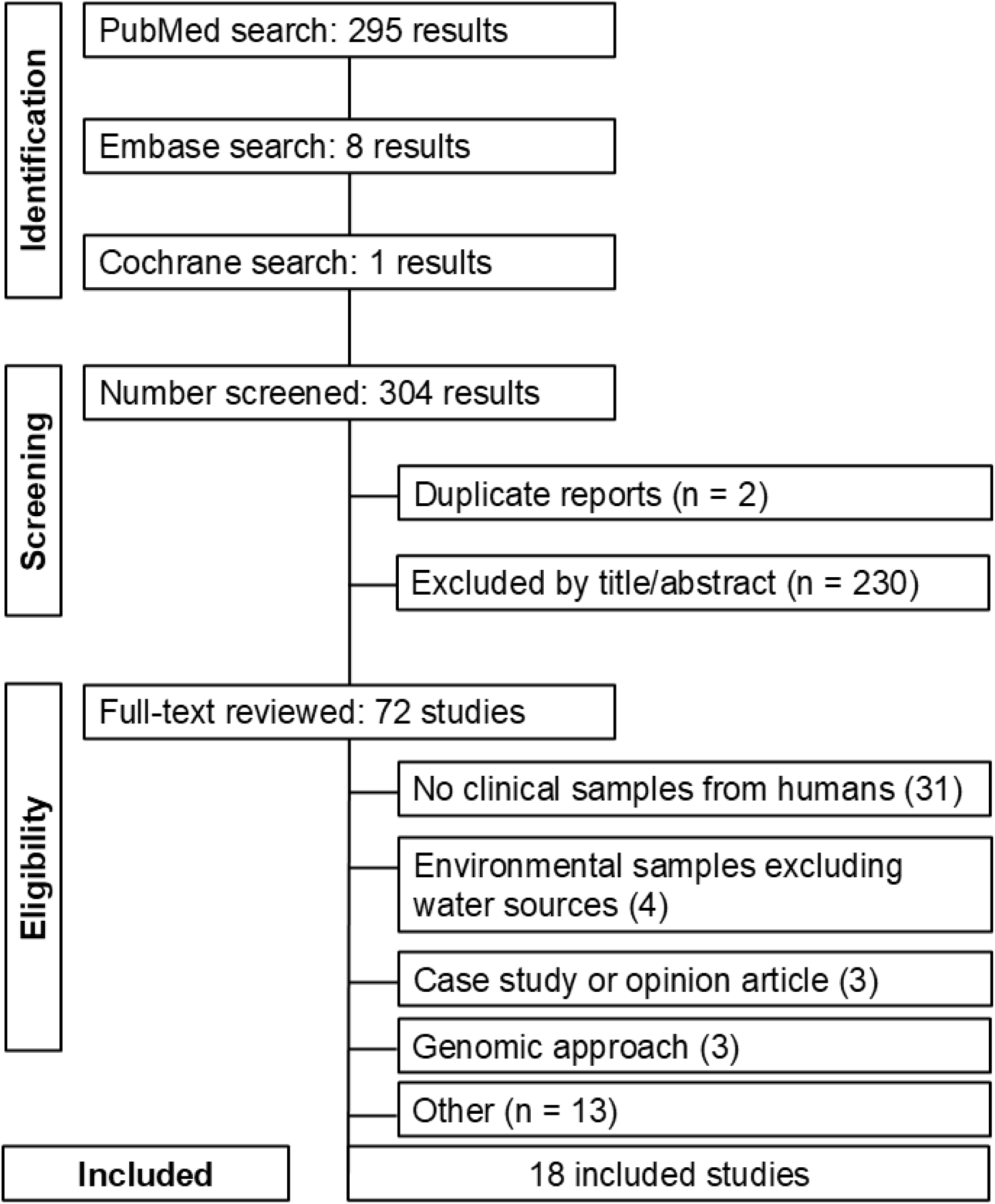
PRISMA flow diagram of study screening and selection.

### 2.2 Eligibility criteria

Included articles were original research published in English reporting AMR in one or more ESKAPE pathogens, with culture-based resistance data from isolates obtained from both environmental matrices (such as water or wastewater) and human clinical samples. Studies were excluded if they were not original research (e.g., reviews, commentaries, editorials, protocols, or case reports without a comparative component), were not focused on ESKAPE pathogens, lacked isolates from either environmental or clinical sources, or relied exclusively on culture-independent approaches without corresponding antimicrobial susceptibility data. Articles not available in full text or lacking a persistent identifier (e.g., DOI) were also excluded. Eligibility criteria were independently applied by two reviewers, with disagreements resolved through discussion and consensus.

### 2.3 Data Sources and Extraction

Data from all eligible studies were systematically extracted by two independent reviewers using a standardized data collection form capturing bibliographic details (first author and year), pathogen, geographic location (country and continent), sample types (environmental and clinical), number of resistant isolates and antibiotics.

For each pathogen, the number of isolates recovered, and the number identified as resistant to specific antimicrobial agents were extracted. Discrepancies between reviewers were resolved through discussion and consensus.

### 2.4 Data analysis

The primary outcome of this meta-analysis was the prevalence of AMR, defined as the proportion of resistant isolates for each specific pathogen-antibiotic combination within each study. For each included study, we extracted the total number of isolates tested, the number of resistant isolates, the specific antibiotic, and the sample source (environmental or clinical). Each unique study–microorganism–antibiotic–source was treated as an independent effect size (Supplementary table 2).

Pooled resistance prevalence was estimated using binomial–normal generalized linear mixed models (GLMMs) with a logit link function [22], accounting for within- and between-study variability. Pooled logit-transformed estimates and corresponding 95% confidence intervals (CIs) were back-transformed to proportions. Between-study heterogeneity was assessed using Cochran’s Q test [23], τ² (maximum likelihood ratio test), and the I² and H² statistics [23]. Homogeneity (τ² = 0) was evaluated using Wald and likelihood-ratio tests. Analyses were conducted for the full dataset and stratified by clinical and environmental sources. Forest plots were generated by antibiotic class, displaying pooled prevalence estimates for clinical and environmental samples, with antibiotics ordered by overall mean resistance. Small-study effects were assessed using contour-enhanced funnel plots of logit-transformed proportions and Egger’s regression test, applied when at least five independent studies were available. All analyses were performed in R (version 4.2.2) [24] using the metafor [25] and meta [26] packages.

## 3. Results

### 3.1 Search and screening results

From this subset, 18 studies met the inclusion criteria and were included in the review. The main reason for exclusion during full-text screening were the absence of phenotypic AMR data, exclusive use of genomic or metagenomic approaches, pre-selection of resistant isolates precluding incidence comparisons, or study scope outside our criteria (e.g., ICU-only or exclusively veterinary samples). One additional study was qualitatively relevant but excluded from the quantitative synthesis due to the lack of isolate incidence data [27]. Consequently, the final meta-analysis was based on data from 18 included studies [28–45].

## 4. Study characteristics

### 4.1 Isolate distribution by pathogen

The included studies varied considerably in scale, with sample sizes ranging from four isolates [42] to over 300 isolates [45]. The majority of studies investigated Enterococcus faecium [28,30,31,34,35,44], Pseudomonas aeruginosa [29,32,33,37,40], or Klebsiella pneumoniae [38,42,43]. In contrast, other ESKAPE pathogens were less frequently represented; Staphylococcus aureus [41], Acinetobacter baumannii [39], and other Klebsiella species [45] were each addressed in a single study. No eligible studies reported isolates belonging to the genus Enterobacter (Supplementary Table 3).

### 4.2 Geographic distribution

The geographical distribution of the included studies is illustrated in Figure 2. Most studies were conducted in Europe, followed by Asia. Africa and Oceania were represented by an equal, smaller number of studies, and North America was the location for a single study. At the national level, Australia was the most frequent study location with three publications. A single study was contributed by each of the following countries: China, Croatia, the Czech Republic, Ethiopia, France, Greece, India, Iran, Italy, Japan, Mexico, Nigeria, Portugal, South Korea, and Spain. The research included in this review covers a period of more than two decades, with the earliest study published in 1999 and the most recent in 2024.

**Figure 2.**
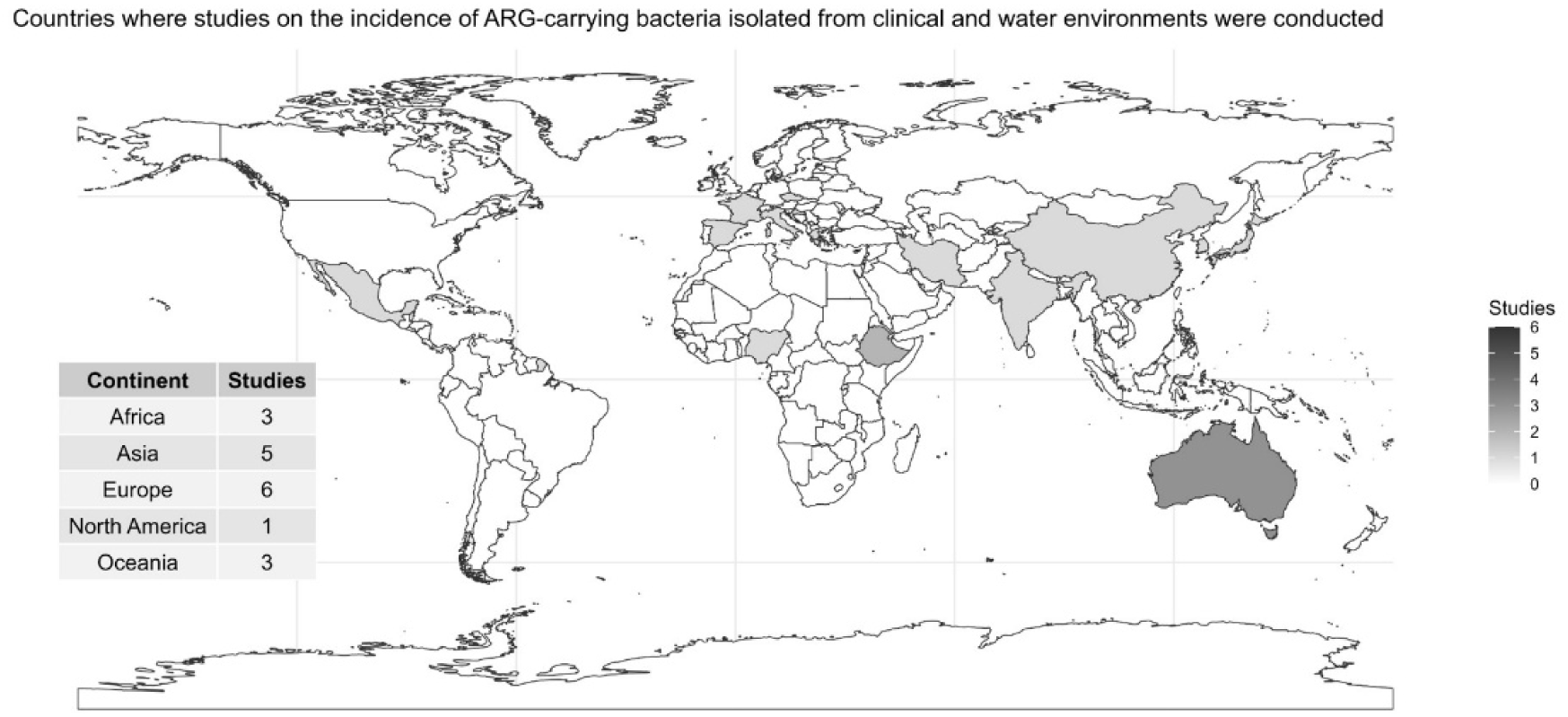
Geographic distribution of research articles investigating antimicrobial resistance in ESKAPE pathogens isolated from clinical and aquatic environments.

### 4.3 Meta-analysis of Antimicrobial Resistance Rates

A meta-analysis was performed to evaluate AMR rates of ESKAPE pathogens across 15 antimicrobial classes and 45 corresponding antibiotic subgroups (Supplementary Figure 1). High heterogeneity was observed across studies (τ² = 14.11; I² = 98.76%; H² = 80.45), as confirmed by Wald and likelihood ratio tests. The overall pooled mean resistance proportion was 0.46 (95% CI: 0.36–0.57).

When stratified by sample origin, heterogeneity remained substantial. Clinical samples showed a pooled mean resistance of 0.67 (95% CI: 0.55–0.77; τ² = 9.62; I² = 97.93%; H² = 48.32), whereas environmental samples exhibited a lower pooled mean resistance of 0.24 (95% CI: 0.14–0.39) with even greater heterogeneity (τ² = 17.24; I² = 99.08%; H² = 108.54) (Supplementary Table 4).

#### 4.3.1 Subgroup analysis by class of antibiotics

A subgroup analysis stratified by sample origin revealed distinct resistance patterns across antibiotic classes. Clinical isolates generally exhibited higher resistance rates than environmental isolates; however, this pattern was reversed for a few classes, including Rifamycins (34.3% environmental vs. 3.3% clinical), Nitrofurans (14.5% vs. 7.5%), Polymyxins (9.5% vs. 3.3%), and Streptogramins (3.4% vs. 2.1%) (Figure 3).

**Figure 3.**
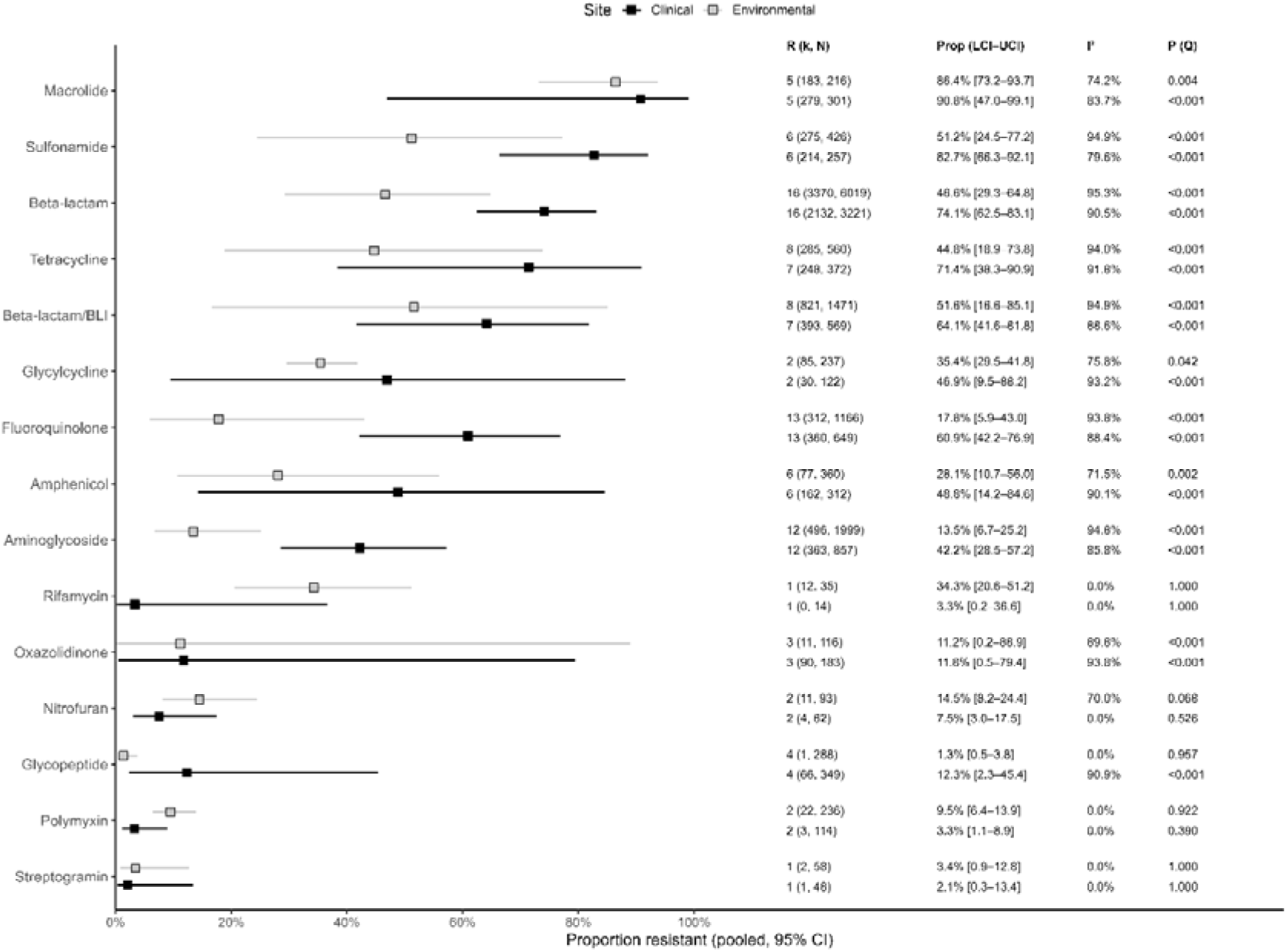
Forest plot of the proportion of antibiotic resistance for the different antibiotic classes.

The highest pooled resistance estimates were observed for Macrolides in both clinical (90.6%) and environmental (86.4%) isolates, followed by Sulfonamides (82.7% vs. 51.2%), β-lactams (74.1% vs. 46.6%), Tetracyclines (71.4% vs. 44.8%), and β-lactam/β-lactamase inhibitor (BL/BLI) combinations (64.1% vs. 51.6%). Intermediate resistance levels were found for Glycylcyclines (46.9% clinical vs. 35.4% environmental), Fluoroquinolones (60.9% vs. 17.8%), Amphenicols (48.8% vs. 28.1%), Aminoglycosides (42.2% vs. 13.5%), and Oxazolidinones, which showed similar rates in both settings (11.8% vs. 11.2%). The lowest resistance proportions were observed for Glycopeptides, with higher values in clinical (12.3%) than in environmental (1.3%) samples (Figure 3).

#### 4.3.2 Subgroup analysis by antibiotics

Analysis at the individual antibiotic level revealed that resistance was generally higher in clinical than in environmental samples for most drugs evaluated (Supplementary Figure 2). The largest discrepancies were observed for ofloxacin, teicoplanin, and vancomycin, with resistance approximately 20-, 14-, and 7-fold higher in clinical samples, respectively. Marked differences were also noted for netilmicin (4.1-fold) and norfloxacin (4-fold).

In contrast, a limited number of antimicrobials exhibited higher resistance in environmental settings, including rifampin (34.3% vs. 3.3% in clinical), cefpodoxime-proxetil (99.3% vs. 34.8%), nitrofurantoin (14.5% vs. 7.5%), colistin (9.5% vs. 3.3%), and quinupristin–dalfopristin (3.4% vs. 2.1%). For several antibiotics (cefuroxime, ampicillin–clavulanic acid, cefoperazone, oxazolidinones, and erythromycin) resistance estimates were similar across both environments.

#### 4.3.3 Subgroup analysis by water matrices

To assess the impact of effluents on AMR, environmental studies were grouped into two subsets: those including at least one effluent-derived source [28,30,31,35,37,39,43,45] and those sampling exclusively from non-effluent waters [29,29,30,34,38,40–42,44]. As most reports did not stratify results by water type, this classification was applied at the study level. Across analyses, between-study heterogeneity remained high (I² ≈ 98%) and statistically significant, indicating true variability among studies. Between-effect variance (τ²) was consistently higher in environmental datasets, with the greatest variance observed in non-effluent waters, suggesting more heterogeneous AMR levels in natural aquatic environments. In contrast, studies including effluents showed lower (τ²), indicating more consistent AMR levels in effluent-impacted settings.

Pooled AMR prevalence was highest in clinical samples compared with both environmental subsets. Within environmental studies, prevalence was significantly higher effluent-including datasets (0.28; 95% CI: 0.17–0.44) than in non-effluent studies (0.15; 95% CI: 0.03–0.49). Conversely, clinical AMR prevalence was lower in studies that also sampled effluents (0.59; 95% CI: 0.58–0.59) than in those without effluent sampling (0.76; 95% CI: 0.63–0.85) (Supplementary Table 4).

#### 4.3.4 Assessment of small-study effects and heterogeneity

Potential small-study effects were assessed using funnel plots and Egger’s regression test for antibiotic classes with at least five available studies, stratified by clinical and environmental samples (Supplementary Figures 3 and Supplementary Table 5). Significant asymmetry was detected for aminoglycosides (t = −2.6, p = 0.02) and fluoroquinolones (t = −3.41, p < 0.001) in environmental samples, and for β-lactams in clinical samples (t = 4.53, p < 0.001) (Supplementary Table 5).

## 5. Discussion

### 5.1 Search and screening results

This systematic review and meta-analysis compared AMR in ESKAPE pathogens from aquatic and clinical sources, identifying 18 eligible studies for inclusion. The limited number of studies highlights a persistent gap in AMR research, particularly the lack of direct comparative analysis integrating environmental and clinical isolates within a One Health framework [7].

Although culture-based approaches capture only a small fraction of environmental microbial diversity [46], they provide direct evidence of phenotypic resistance by demonstrating the survival of viable bacteria under antimicrobial exposure. This functional confirmation allows a more reliable assessment of active resistance potential across environmental and clinical compartments [47].

### 5.2 Study characteristics

#### 5.2.1 Isolate distribution by pathogen

In this review, the most frequently reported ESKAPE pathogens were *E. faecium*, *P. aeruginosa*, and *K. pneumoniae*, all well-documented opportunistic pathogens associated with multidrug resistance and healthcare-associated infections [13,48]. Their predominance in studies comparing clinical and environmental samples likely reflects their ecological adaptability, allowing persistence across diverse niches and facilitating recovery under laboratory conditions. *E. faecium* is a ubiquitous organism found in sources ranging from animal gastrointestinal tracts to water, soil, and wastewater, largely due to its tolerance to adverse conditions such as variations in temperature, salinity, and pH [12,13]. Similarly, *P. aeruginosa* is an ecologically versatile species common in water and soil, whose metabolic flexibility and capacity to form biofilms support persistence in both natural and hospital ecosystems [49]. *K. pneumoniae* also occupied diverse niches, being frequently detected in water and soil and capable of surviving under both aerobic and anaerobic conditions [14].

In contrast, *S. aureus* and *A. baumannii* exhibit narrower ecological ranges. *S. aureus* is primarily adapted to colonize human skin and mucosa and is typically detected at lower concentrations in environmental samples [15,50], whereas *A. baumannii* remains predominantly associated with hospital environments, despite occasional recovery from soil or water [51].

No eligible studies reported *Enterobacter* species. Although members of this genus have been detected in soil, wastewater, and aquatic ecosystems [16], their absence may reflect lower prevalence in human infections compared to other ESKAPE pathogens [52], as well as challenges in species-level identification using conventional methods.

#### 5.2.2 Geographic distribution

The geographic distribution of the included studies indicates a marked bias in AMR research on ESKAPE pathogens, with most studies conducted in high-income countries. This pattern likely reflects differences in research infrastructure, funding availability, and maturity of AMR surveillance programs. Such disparities are captured by the Global One Health Index for AMR (GOHI-AMR), which consistently shows higher scores for high-income countries [53], reinforcing the impact of unequal monitoring capacity and research development on the global AMR evidence base.

### 5.3 Meta-analysis

#### 5.3.1 Sources of Inter-Study Heterogeneity

High between-study heterogeneity persisted even after dataset stratifying, indicating that the observed variability reflects genuine methodological and ecological differences rather than sampling error. Environmental studies retained substantial heterogeneity even when divided into effluent and non-effluent subsets, largely driven by diversity in sample matrices and variations in laboratory methods, which directly influence observed resistance proportions. Ecological Heterogeneity: The Role of Sample Matrix

A major driver of variability was the pooling of multiple, ecologically distinct sample types within single studies. For example, Giri et al. [43] combined residential well water and fish-market effluent, while Pinto et al. [28] analyzed samples ranging from compost and swimming pools to marine water and wastewater. Aggregating such heterogeneous matrices introduces substantial ecological noise, as each environment is subject to distinct selective pressure. In contrast, studies focusing exclusively on wastewater matrices [31,39] may reduce this variability but often still report high resistance levels. Wastewater represents a well-established hotspot, enriched with antimicrobial residues, metals, and dense microbial communities that promote the selection and dissemination of resistance genes, thereby elevating observed resistance proportions [54].

#### 5.3.2 Methodological Heterogeneity: The Impact of Isolation Techniques

A second major source of variability was the laboratory methodology used for bacterial isolation. Some studies employed antibiotic-supplemented media to enhance recovery of resistant isolates such as the use of gentamicin-, cefotaxime-, or meropenem-containing media [31,45]. Although effective for targeting resistant strains, this pre-selection approach inflates resistance estimates compared with studies using non-selective media. In contrast, other investigations relied on standard differential media, including MacConkey agar [43] or cetrimide agar [37], which do not artificially enrich resistant subpopulations. These differences in isolation strategies limit direct cross-study comparisons and substantially contribute to the heterogeneity observed in this meta-analysis (Supplementary table 3).

#### 5.3.3 Subgroup analysis by class of antibiotics and antibiotics Higher Resistance in Clinical Isolates

The pooled prevalence of resistant ESKAPE isolates was consistently higher in clinical samples than in environmental ones, even after stratifying environmental data into effluent and non-effluent subsets. This finding is consistent with large-scale clinical syntheses showing a marked temporal escalation of antimicrobial resistance in ESKAPE pathogens; notably, a nationwide review from India documented a substantial increase in resistance rates across multiple antibiotic classes over the 2010–2020 decade, with the highest resistance levels observed in the latter half of the period [55]. This pattern reflects the strong selective pressures typical of healthcare settings, including intensive antibiotic use, frequent invasive procedures, high patient density, and the concentration of vulnerable populations, all of which favor the selection and dissemination of resistant strains [3].

##### Higher Environmental Resistance for Four Antibiotic Classes

In contrast, rifamycins, nitrofurans, polymyxins, and streptogramins showed higher pooled resistance in environmental isolates. However, this pattern does not appear to represent a true ecological signal but rather an artifact driven by specific study contexts and methodologies. The studies contributing most to this finding primarily sampled wastewater from a limited number of locations (Mexico [35], Iran [31], and the Czech Republic [45]). The sole exception was a study from Australia [42], which analyzed river water but was based on a very small sample size. Importantly, several of these studies employed selective isolation approaches, including pre-selection for high-level gentamicin resistance [31] of the use of antibiotic-supplemented media [45], which artificially enrich resistant phenotypes and inflate resistance estimates in environmental samples, thereby confounding direct comparisons with clinical isolates.

#### 5.3.4 Small-Study Effects Driven by Uneven Antibiotic Representation

Significant funnel plot asymmetry was observed for aminoglycosides and fluoroquinolones in environmental samples and for β-lactams in clinical samples. Although such asymmetry is often interpreted as publication bias, our findings indicate that it is more plausibly explained by methodological heterogeneity, particularly the uneven representation of individual antibiotics within each aggregated class. For environmental aminoglycosides (resistance range: 5.2%–65.6%), netilmicin data were derived from only two studies [30,45], whereas gentamicin was assessed in twelve [29,30,32–35,37,39,41–43,45]. A similar imbalance was observed for fluoroquinolones (range: 0.6%–83.7%), with norfloxacin evaluated in only two studies [30,37] and ciprofloxacin in eleven [30,31,33,34,37–39,42–45]. The negative t-values indicate that smaller studies tended to report higher resistance, likely reflecting the influence of sparsely tested antibiotics with high resistance estimates.

An inverse pattern was observed for clinical β-lactams (range: 11.3%–99.3%), where smaller studies reported lower resistance. This is consistent with aggregation of rarely tested agents such as cefpodoxime-proxetil, assessed in a single study [45], with widely tested antibiotics imipenem, included in ten studies [28–31,33–35,42,43,45].

Overall, these small-study effects do not reflect classic publication bias but arise from the aggregation of heterogeneous data at the antibiotic-class level. This underscores a key methodological challenge in AMR meta-analyses: uneven representation of individual drugs can distort heterogeneity assessments and generate misleading statistical signals.

## 6. Conclusion

This meta-analysis reveals a complex and highly heterogeneous landscape of AMR in ESKAPE pathogens across clinical and environmental settings. Our findings confirm that resistance prevalence is significantly higher in clinical isolates, reflecting the intense selective pressures in healthcare environments characterized by high antibiotic use and vulnerable patient populations. However, this overall pattern is strongly influenced by substantial methodological and ecological heterogeneity among studies. This high statistical heterogeneity observed was not merely a sampling artifact but a genuine reflection of variability in the primary literature, driven by the pooling of distinct environmental matrices and the lack of standardized laboratory protocols.

These limitations were most evident in the anomalous finding that rifamycins, nitrofurans, polymyxins, and streptogramins showed higher resistance in environmental samples. This counterintuitive result did not represent a true ecological signal but rather an artifact arising from a small number of studies focused on wastewater hotspots and employing enrichment techniques that inflate resistance estimates. Consistently, Egger’s test indicated that funnel plot asymmetry for aminoglycosides, fluoroquinolones, and β-lactams was not indicative of publication bias but instead reflected uneven representation of individual antibiotics within aggregated classes.

In summary, although antimicrobial resistance remains more concentrated in clinical settings, our synthesis highlights a critical limitation in current AMR research: the lack of standardized sampling and analytical frameworks severely constrains data integration. The contribution of environmental reservoirs to the clinical AMR burden cannot be accurately quantified until harmonized methodologies are adopted. Future studies should prioritize standardized protocols to improve comparability and enable a clearer understanding of transmission dynamics between environmental and clinical compartments.

## Supporting information

Search terms

Characteristics of included studies and antimicrobial resistance profiles of bacterial isolates from clinical and environmental sources

Overview of inlcuded studies

. Pooled antimicrobial resistance and heterogeneity metrics by site and dataset type

Publication heterogeneity on the patterns of antimicrobial resistance in clinical and environmental samples

Comparison of pooled antimicrobial resistance proportions between clinical and environmental bacterial isolates by antimicrobial class and agent

Forest plots of pooled antimicrobial resistance proportions by antibiotic class comparing clinical and environmental bacterial isolates

Funnel plots of pooled antimicrobial resistance estimates for clinical and environmental isolates by antibiotic class

PRISMA checklist

## Data Availability

All data produced in the present study are available upon reasonable request to the authors

## Author contribution

A. B. M. Vaz: Conceptualization, Data curation, Formal analysis, Investigation, Methodology, Project administration, Writing – original draft

B. Murad: Conceptualization, Data curation

B. Lopes: Conceptualization, Data curation

M. L. Castro: Conceptualization, Data curation

G. Fernandes: Conceptualization, Writing – review and editing

W. K. Oliveira: Conceptualization, Methodology

P. L. C. Fonseca: Writing – review and editing

E. R. G. R. Aguiar: Writing – review and editing

C. R. Mota: Writing – review and editing

A. B. Santos: Writing – review and editing

C. Starling: Conceptualization, Funding acquisition, Investigation, Methodology, Project administration, Supervision, Writing – review and editing.

## Declaration of generative AI use

During the preparation of this work, the author(s) used Resea in order to improve the clarity, conciseness, and overall readability of the manuscript. After using this service, the author(s) reviewed and edited the content as needed and took full responsibility for the content of the published article.

## Supplementary files

**Supplementary Figure 1**. Comparison of pooled antimicrobial resistance proportions between clinical and environmental bacterial isolates by antimicrobial class and agent.

**Supplementary Figure 2**. Forest plots of pooled antimicrobial resistance proportions by antibiotic class comparing clinical and environmental bacterial isolates.

**Supplementary Figure 3**. Funnel plots of pooled antimicrobial resistance estimates for clinical and environmental isolates by antibiotic class.

**Supplementary table 1**. Search terms

**Supplementary table 2**. Characteristics of included studies and antimicrobial resistance profiles of bacterial isolates from clinical and environmental sources.

**Supplementary table 3**. Overview of clinical and environmental samples, sample sources, number of isolates, geographic distribution, ESKAPE pathogens, isolation methods and identification techniques in the included studies.

**Supplementary table 4**. Pooled antimicrobial resistance and heterogeneity metrics by site (clinical and environmental) and dataset type (all, with effluent, without effluent.

**Supplementary table 5**. Publication heterogeneity on the patterns of antimicrobial resistance in clinical and environmental samples.

## References

[1] Brooks G, Carroll KC, Butel J, Morse S. Lange Medical Microbiology 2007.

[2] Dadgostar P. Antimicrobial Resistance: Implications and Costs. Infect Drug Resist 2019;12:3903–10. 10.2147/IDR.S234610.

[3] Prestinaci F, Pezzotti P, Pantosti A. Antimicrobial resistance: a global multifaceted phenomenon. Pathog Glob Health 2015;109:309–18. 10.1179/2047773215Y.0000000030.

[4] Ho CS, Wong CTH, Aung TT, Lakshminarayanan R, Mehta JS, Rauz S, et al. Antimicrobial resistance: a concise update. Lancet Microbe 2025;6:100947. 10.1016/j.lanmic.2024.07.010.

[5] Ching C, Orubu ESF, Sutradhar I, Wirtz VJ, Boucher HW, Zaman MH. Bacterial antibiotic resistance development and mutagenesis following exposure to subinhibitory concentrations of fluoroquinolones in vitro: a systematic review of the literature. JAC Antimicrob Resist 2020;2:dlaa068. 10.1093/jacamr/dlaa068.

[6] Queenan K, Häsler B, Rushton J. A One Health approach to antimicrobial resistance surveillance: is there a business case for it? Int J Antimicrob Agents 2016;48:422–7. 10.1016/j.ijantimicag.2016.06.014.

[7] Hernando-Amado S, Coque TM, Baquero F, Martínez JL. Defining and combating antibiotic resistance from One Health and Global Health perspectives. Nat Microbiol 2019;4:1432–42. 10.1038/s41564-019-0503-9.

[8] World Health Organization. AWaRe classification of antibiotics for evaluation and monitoring of use, 2023. World Health Organization: Geneva, Switzerland 2023.

[9] Rice LB. Federal funding for the study of antimicrobial resistance in nosocomial pathogens: no ESKAPE. J Infect Dis 2008;197:1079–81. 10.1086/533452.

[10] Pendleton JN, Gorman SP, Gilmore BF. Clinical relevance of the ESKAPE pathogens. Expert Rev Anti Infect Ther 2013;11:297–308. 10.1586/eri.13.12.

[11] Santajit S, Indrawattana N. Mechanisms of Antimicrobial Resistance in ESKAPE Pathogens. Biomed Res Int 2016;2016:2475067. 10.1155/2016/2475067.

[12] Fisher K, Phillips C. The ecology, epidemiology and virulence of Enterococcus. Microbiology (Reading) 2009;155:1749–57. 10.1099/mic.0.026385-0.

[13] Denissen J, Reyneke B, Waso-Reyneke M, Havenga B, Barnard T, Khan S, et al. Prevalence of ESKAPE pathogens in the environment: Antibiotic resistance status, community-acquired infection and risk to human health. Int J Hyg Environ Health 2022;244:114006. 10.1016/j.ijheh.2022.114006.

[14] Araújo S, Silva V, Quintelas M, Martins Â, Igrejas G, Poeta P. From soil to surface water: exploring Klebsiella’s clonal lineages and antibiotic resistance odyssey in environmental health. BMC Microbiol 2025;25:97. 10.1186/s12866-025-03798-8.

[15] Goodwin KD, McNay M, Cao Y, Ebentier D, Madison M, Griffith JF. A multi-beach study of Staphylococcus aureus, MRSA, and enterococci in seawater and beach sand. Water Res 2012;46:4195–207. 10.1016/j.watres.2012.04.001.

[16] Davin-Regli A, Lavigne J-P, Pagès J-M. Enterobacter spp.: Update on Taxonomy, Clinical Aspects, and Emerging Antimicrobial Resistance. Clin Microbiol Rev 2019;32:e00002–19. 10.1128/CMR.00002-19.

[17] Cycoń M, Mrozik A, Piotrowska-Seget Z. Antibiotics in the Soil Environment-Degradation and Their Impact on Microbial Activity and Diversity. Front Microbiol 2019;10:338. 10.3389/fmicb.2019.00338.

[18] Berendonk TU, Manaia CM, Merlin C, Fatta-Kassinos D, Cytryn E, Walsh F, et al. Tackling antibiotic resistance: the environmental framework. Nat Rev Microbiol 2015;13:310–7. 10.1038/nrmicro3439.

[19] Wellington EMH, Boxall AB, Cross P, Feil EJ, Gaze WH, Hawkey PM, et al. The role of the natural environment in the emergence of antibiotic resistance in gram-negative bacteria. Lancet Infect Dis 2013;13:155–65. 10.1016/S1473-3099(12)70317-1.

[20] Moher D, Liberati A, Tetzlaff J, Altman DG. Preferred reporting items for systematic reviews and meta-analyses: the PRISMA Statement. Open Med 2009;3:e123–130.

[21] Page MJ, McKenzie JE, Bossuyt PM, Boutron I, Hoffmann TC, Mulrow CD, et al. The PRISMA 2020 statement: an updated guideline for reporting systematic reviews. Bmj 2021;372.

[22] Harrer M, Cuijpers P, Furukawa T, Ebert D. Doing meta-analysis with R: A hands-on guide. Chapman and Hall/CRC; 2021.

[23] Lin L, Chu H. Meta-analysis of proportions using generalized linear mixed models. Epidemiology 2020;31:713–7.

[24] R Core Team. R: A Language and Environment for Statistical Computing 2022.

[25] Viechtbauer W. Conducting Meta-Analyses in R with the metafor Package. J Stat Soft 2010;36. 10.18637/jss.v036.i03.

[26] Balduzzi S, Rücker G, Schwarzer G. How to perform a meta-analysis with R: a practical tutorial. Evid Based Mental Health 2019;22:153–60. 10.1136/ebmental-2019-300117.

[27] Varela AR, Macedo GN, Nunes OC, Manaia CM. Genetic characterization of fluoroquinolone resistant Escherichia coli from urban streams and municipal and hospital effluents. FEMS Microbiol Ecol 2015;91:fiv015. 10.1093/femsec/fiv015.

[28] Pinto B, Pierotti R, Canale G, Reali D. Characterization of “faecal streptococci” as indicators of faecal pollution and distribution in the environment. Lett Appl Microbiol 1999;29:258–63. 10.1046/j.1472-765x.1999.00633.x.

[29] Khan NH, Ishii Y, Kimata-Kino N, Esaki H, Nishino T, Nishimura M, et al. Isolation of Pseudomonas aeruginosa from open ocean and comparison with freshwater, clinical, and animal isolates. Microb Ecol 2007;53:173–86. 10.1007/s00248-006-9059-3.

[30] Kotzamanidis C, Zdragas A, Kourelis A, Moraitou E, Papa A, Yiantzi V, et al. Characterization of vanA-type Enterococcus faecium isolates from urban and hospital wastewater and pigs. J Appl Microbiol 2009;107:997–1005. 10.1111/j.1365-2672.2009.04274.x.

[31] Saifi M, Pourshafie MR, Dallal MMS, Katouli M. Clonal groups of high-level gentamicin-resistant Enterococcus faecium isolated from municipal wastewater and clinical samples in Tehran, Iran. Lett Appl Microbiol 2009;49:160–5. 10.1111/j.1472-765X.2009.02559.x.

[32] Kim JR, Lee DK, An HM, Kim MJ, Lee SW, Cha MK, et al. Antimicrobial activity of commonly used antibiotics and DNA fingerprint analysis of Pseudomonas aeruginosa obtained from clinical isolates and unchlorinated drinking water in Korea, 2010. Arch Pharm Res 2011;34:1353–61. 10.1007/s12272-011-0816-6.

[33] Ruiz-Martínez L, López-Jiménez L, Fusté E, Vinuesa T, Martínez JP, Viñas M. Class 1 integrons in environmental and clinical isolates of Pseudomonas aeruginosa. Int J Antimicrob Agents 2011;38:398–402. 10.1016/j.ijantimicag.2011.06.016.

[34] Rathnayake IU, Hargreaves M, Huygens F. Antibiotic resistance and virulence traits in clinical and environmental Enterococcus faecalis and Enterococcus faecium isolates. Syst Appl Microbiol 2012;35:326–33. 10.1016/j.syapm.2012.05.004.

[35] Castillo-Rojas G, Mazari-Hiríart M, Ponce de León S, Amieva-Fernández RI, Agis-Juárez RA, Huebner J, et al. Comparison of Enterococcus faecium and Enterococcus faecalis Strains isolated from water and clinical samples: antimicrobial susceptibility and genetic relationships. PLoS One 2013;8:e59491. 10.1371/journal.pone.0059491.

[36] Varela AR, Ferro G, Vredenburg J, Yanık M, Vieira L, Rizzo L, et al. Vancomycin resistant enterococci: from the hospital effluent to the urban wastewater treatment plant. Sci Total Environ 2013;450–451:155–61. 10.1016/j.scitotenv.2013.02.015.

[37] Streeter K, Neuman C, Thompson J, Hatje E, Katouli M. The characteristics of genetically related Pseudomonas aeruginosa from diverse sources and their interaction with human cell lines. Can J Microbiol 2016;62:233–40. 10.1139/cjm-2015-0536.

[38] Abera B, Kibret M, Mulu W. Extended-Spectrum beta (β)-Lactamases and Antibiogram in Enterobacteriaceae from Clinical and Drinking Water Sources from Bahir Dar City, Ethiopia. PLoS One 2016;11:e0166519. 10.1371/journal.pone.0166519.

[39] Kovacic A, Seruga Music M, Dekic S, Tonkic M, Novak A, Rubic Z, et al. Transmission and survival of carbapenem-resistant Acinetobacter baumannii outside hospital setting. Int Microbiol 2017;20:165–9. 10.2436/20.1501.01.299.

[40] Tran-Dinh A, Neulier C, Amara M, Nebot N, Troché G, Breton N, et al. Impact of intensive care unit relocation and role of tap water on an outbreak of Pseudomonas aeruginosa expressing OprD-mediated resistance to imipenem. J Hosp Infect 2018;100:e105–14. 10.1016/j.jhin.2018.05.016.

[41] Adesoji AT, Olatoye IO, Ogunjobi AA. Genotypic Characterization of Aminoglycoside Resistance Genes from Bacteria Isolates in Selected Municipal Drinking Water Distribution Sources in Southwestern Nigeria. Ethiop J Health Sci 2019;29:321–32. 10.4314/ejhs.v29i3.4.

[42] Lepuschitz S, Schill S, Stoeger A, Pekard-Amenitsch S, Huhulescu S, Inreiter N, et al. Whole genome sequencing reveals resemblance between ESBL-producing and carbapenem resistant Klebsiella pneumoniae isolates from Austrian rivers and clinical isolates from hospitals. Sci Total Environ 2019;662:227–35. 10.1016/j.scitotenv.2019.01.179.

[43] Giri S, Shekar M, Shetty AV, G PT, Shetty AK. Antibiotic resistance and random amplified polymorphic DNA typing of Klebsiella pneumoniae isolated from clinical and water samples. Water Environ Res 2021;93:2740–53. 10.1002/wer.1630.

[44] Shen W, Cai C, Dong N, Chen J, Zhang R, Cai J. Mapping the widespread distribution and transmission dynamics of linezolid resistance in humans, animals, and the environment. 2024;12.

[45] Davidova-Gerzova L, Lausova J, Sukkar I, Nechutna L, Kubackova P, Krutova M, et al. Multidrug-resistant ESBL-producing Klebsiella pneumoniae complex in Czech hospitals, wastewaters and surface waters. Antimicrob Resist Infect Control 2024;13:141. 10.1186/s13756-024-01496-0.

[46] Staley JT, Konopka A. Measurement of in situ activities of nonphotosynthetic microorganisms in aquatic and terrestrial habitats. Annu Rev Microbiol 1985;39:321–46. 10.1146/annurev.mi.39.100185.001541.

[47] Hsu C-Y, Moradkasani S, Suliman M, Uthirapathy S, Zwamel AH, Hjazi A, et al. Global patterns of antibiotic resistance in group B Streptococcus: a systematic review and meta-analysis. Front Microbiol 2025;16:1541524. 10.3389/fmicb.2025.1541524.

[48] Denissen J, Havenga B, Reyneke B, Khan S, Khan W. Comparing antibiotic resistance and virulence profiles of Enterococcus faecium, Klebsiella pneumoniae, and Pseudomonas aeruginosa from environmental and clinical settings. Heliyon 2024;10:e30215. 10.1016/j.heliyon.2024.e30215.

[49] Schroth MN, Cho JJ, Green SK, Kominos SD, Publishing MS. Epidemiology of Pseudomonas aeruginosa in agricultural areas(). J Med Microbiol 2018;67:1191–201. 10.1099/jmm.0.000758.

[50] Tong SYC, Davis JS, Eichenberger E, Holland TL, Fowler VGJ. Staphylococcus aureus infections: epidemiology, pathophysiology, clinical manifestations, and management. Clin Microbiol Rev 2015;28:603–61. 10.1128/CMR.00134-14.

[51] Peleg AY, Seifert H, Paterson DL. Acinetobacter baumannii: emergence of a successful pathogen. Clinical Microbiology Reviews 2008;21:538–82.

[52] Khasapane NG, Nkhebenyane SJ, Lekota K, Thekisoe O, Ramatla T. “One Health” Perspective on Prevalence of ESKAPE Pathogens in Africa: A Systematic Review and Meta-Analysis. Pathogens 2024;13. 10.3390/pathogens13090787.

[53] Zhou H, Xu X, Wangjin Y, Ye M, Wu T, Wang Z, et al. Degradation of sulfamethoxazole and antibiotic resistance genes from surface water in the photocatalyst-loading bionic ecosystems. Sci Total Environ 2023;895:165045. 10.1016/j.scitotenv.2023.165045.

[54] Kunhikannan S, Thomas CJ, Franks AE, Mahadevaiah S, Kumar S, Petrovski S. Environmental hotspots for antibiotic resistance genes. Microbiologyopen 2021;10:e1197.

[55] Kharat AS, Makwana N, Nasser M, Gayen S, Yadav B, Kumar D, et al. Dramatic increase in antimicrobial resistance in ESKAPE clinical isolates over the 2010–2020 decade in India. International Journal of Antimicrobial Agents 2024;63:107125. 10.1016/j.ijantimicag.2024.107125.

