## Supplementary material for "Antimicrobial resistance prevalence in clinical and aquatic environmental ESKAPE: a systematic review with meta-analysis": Search terms

**Supplementary table 1.** Search terms

| | |
|---|---|
| Pubmed | ("Antibiotic Resistance"[MeSH] OR "Drug Resistance, Microbial"[MeSH] OR ARG OR Resistome) AND ("Anti-Bacterial Agents"[MeSH] OR Azithromycin OR Ciprofloxacin OR Clindamycin OR Norfloxacin OR Carbapenem) AND (“Enterobacter” OR "Enterococcus" OR "Staphylococcus" OR "Pseudomonas" OR "Klebsiella" OR "Acinetobacter") AND ("Water"[MeSH] OR "Environmental Microbiology"[MeSH] OR River OR Lake OR Ocean) and (Clinical infection) |
| Embase | (("Antibiotic Resistance" OR "Drug Resistance, Microbial" OR ARG OR Resistome) AND ("Anti-Bacterial Agents" OR Azithromycin OR Ciprofloxacin OR Clindamycin OR Norfloxacin OR Carbapenem) AND (“Enterobacter” OR "Enterococcus" OR "Staphylococcus" OR "Pseudomonas" OR "Klebsiella" OR "Acinetobacter") AND ("Water" OR "Environmental Microbiology" OR River OR Lake OR Ocean)) AND (Clinical infection) |
| Cochrane | ("Antibiotic Resistance" OR "Drug Resistance, Microbial" OR ARG OR Resistome) AND ("Anti-Bacterial Agents" OR Azithromycin OR Ciprofloxacin OR Clindamycin OR Norfloxacin OR Carbapenem) AND (“Enterobacter” OR "Enterococcus" OR "Staphylococcus" OR "Pseudomonas" OR "Klebsiella" OR "Acinetobacter") AND ("Water" OR "Environmental Microbiology" OR River OR Lake OR Ocean) AND (Clinical infection) |
