## Supplementary material for "Antimicrobial resistance prevalence in clinical and aquatic environmental ESKAPE: a systematic review with meta-analysis": Overview of inlcuded studies

**Supplementary table 3.** Overview of clinical and environmental samples, sample sources, number of isolates, geographic distribution, ESKAPE pathogens, isolation methods and identification techniques in the included studies.

| Article identification | Types of samples | Number of isolates | Country | Targeted ESKAPE pathogen(s) | Isolation and presumptive identification | Typing methods and Identification |
|---|---|---|---|---|---|---|
| 28 | **Clinical**<br>- Urine<br><br>**Environmental**<br>- Domestic refuse composting<br>- Public swimming pool water<br>- Marine brackish<br>- Effluent<br>- Sediments from harbour dredging activity | Clinical: 57<br>Environmental: 59 | Italy | *Enterococcus faecium* | **Clinical**<br>The isolates were obtained from the clinical microbiology department.<br><br>**Environmental**<br>**Methodology: Most probable number (MPN) method**<br>- Azide Dextrose Broth [S]<br>- Esculin vancomycin azide broth [S]<br>- Bile Esculin Agar [S, D]<br>- Catalase<br>- Gram-staining | **Clinical**<br>**Methodology: Biochemical identification**<br>- Automated: SCEPTOR system<br><br>**Environmental**<br>**Methodology: Presumptive phenotypic and Biochemical identification**<br>Manual and automated identification (SCEPTOR system) |
| 29 | **Clinical**<br>- Blood, urine, sputum, pus, cow, rabbit, wild boar<br><br>**Environmental**<br>- River water<br>- Lake, ponds and marine water<br>- Surface seawater | Clinical: 68<br>Environmental: 23 | Japan | *Pseudomonas aeruginosa* | **Clinical**<br>The isolates were previously isolated<br><br>**Environmental**<br>**Methodology: Membrane filtration and culture**<br>- Nalidixic acid cetrimide (NAC) agar [S]<br>- Nutrient broth agar (NA) [E]<br>Cetrimide kanamycin nalidixic acid agar [S] | **Clinical and environmental**<br>**Methodology: Biochemical and Genotypic identification**<br>- Automated: BD Phoenix Automated microbiology system<br>- Serotyping<br>PFGE |
| 30 | **Clinical**<br>- Blood cultures of patients hospitalized<br><br>**Environmental**<br>- Wastewater (raw)<br>- Effluent (treated)<br>- Secondary treated and disinfected wastewater from effluents of | Clinical: 10<br>Environmental: 53 | Greece | *Enterococcus faecium* | **Clinical**<br>**Methodology: Biochemical and culture**<br>- Automated system Bactec 9240<br>- Blood agar [E]<br><br>**Environmental**<br>**Methodology: Direct plating**<br>- Kanamycin aesculin azide agar [S] supplemented with | **Clinical**<br>**Methodology: Biochemical and Molecular fingerprint identification**<br>- Automated: API Strep test<br>- Automated: VITEK2 automated system<br><br>**Environmental**<br>**Methodology: Biochemical and Molecular fingerprint identification** |

| | | | | | | |
|---|---|---|---|---|---|---|
| | hospitals<br>- Pig sample (rectum and faeces) | | | | vancomycin<br>- Catalase<br>- Gram-staining<br>Motility | - Manual: Biochemical<br>PFGE |
| 31 | **Clinical**<br>- Urine and wound<br><br>**Environmental**<br>- Wastewater (raw) | Clinical: 48<br>Environmental: 58 | Iran | *Enterococcus faecium* | **Clinical**<br>It was not described<br><br>**Environmental**<br>**Methodology: Membrane filtration and culture**<br>- Brain heart infusion (BHI) agar [E]<br>m-*Enterococcus* agar [S] supplemented with gentamicin. | **Clinical and environmental**<br>**Methodology: Genotypic and phenotypic identification**<br>- PCR<br>- PhP-RF system<br>- Ribotyping<br>- PFGE |
| 32 | **Clinical**<br>- Pus, urine, sputum, bronchial wash solution<br><br>**Environmental**<br>- Spring water | Clinical: 6<br>Environmental: 12 | South Korea | *Pseudomonas aeruginosa* | **Clinical and Environmental**<br><br>**Methodology: Direct plating**<br>- *Pseudomonas* isolation agar [S] | **Clinical and Environmental**<br><br>**Methodology: Presumptive phenotypic and Molecular fingerprint identification**<br>RAPD |
| 33 | **Clinical**<br>- Clinical samples (it was not specified)<br><br>**Environmental**<br>- Water (tap water)<br>- Garden | Clinical: 56<br>Environmental: 20 | Spain | *Pseudomonas aeruginosa* | **Clinical and Environmental**<br>**Methodology: Biochemical and Culture**<br>- Trypticase soy broth (TSB) [E]<br>- Glutamate starch Pseudomonas agar (GSP) [S, D]<br>- Gram staining<br>- Positive oxidase reaction<br>- O/F +/- | **Clinical and Environmental**<br>**Methodology: Phenotypic and genotypic identification**<br>- SDS-PAGE<br>- PFGE<br>RAPD |
| 34 | **Clinical**<br>- Obtained from Pathology Queensland and QUT culture collection [did not specify the sample site origin]<br><br>**Environmental**<br>- River water | Clinical: 27<br>Environmental: 47 | Australi a | *Enterococcus faecium* | **Clinical**<br>The isolated were obtained from culture collection<br><br>**Environmental**<br>**Methodology: Membrane filtration and culture**<br>- *Enterococcus* agar [S] | **Clinical**<br>The bacterial species were identified according to the classification provided by the reference culture collection, since the isolates were sourced directly from it.<br><br>**Environmental**<br><br>**Methodology: Presumptive phenotypic and Genotypic identification**<br>Real-time PCR using $ddl_{E.\ faecalis}$ and $ddl_{E.feacium}$ |

| | | | | | | |
|---|---|---|---|---|---|---|
| 35 | **Clinical**<br>- Blood, cerebrospinal fluid, eyes, livers, peripheral venous catheters, pleural fluid, sputum, urine, wounds, respiratory system<br><br>**Environmental**<br>- Groundwater for human use (wells)<br>- Wetland (lake)<br>- Wastewater (treated) | Clinical: 14<br>Environmental: 35 | Mexico | *Enterococcus faecium, Enterococcus faecalis* | **Clinical**<br>**Method: Culture**<br>- Sheep blood agar [E]<br>- Grain-staining<br>- Catalase<br>- Esculin hydrolysis<br>- BHI 6.5% NaCl<br><br>**Environmental**<br>**Method: Membrane filtration and culture**<br>- K-F agar [S, D]<br>- Grain-staining<br>- Catalase<br>- Esculin hydrolysis<br>- BHI 6.5% NaCl | **Clinical**<br>**Methodology: Genotypic and phenotypic identification**<br>- Multiplex PCR<br>PFGE |
| 37 | **Clinical**<br>- Hospitalized patients: urinary and respiratory tract infections<br><br>**Environmental**<br>- Wastewater (raw)<br>- River water | Clinical: 20<br>Environmental: 214 | Australi a | *Pseudomonas aeruginosa* | **Clinical**<br>**Methodology: Culture**<br>- Previously isolated by the pathology unit of the hospital<br><br>**Environmental**<br>**Methodology: Membrane filtration and culture**<br>Cetrimide agar [S] | **Clinical and Environmental**<br>**Genotypic fingerprinting and molecular detection**<br>- PCR<br>RAPD |
| 38 | **Clinical**<br>- Blood<br><br>**Environmental**<br>- Tap water | Clinical: 49<br>Environmental: 6 | Ethiopi a | *Klebsiella pneumoniae* | **Clinical**<br>**Methodology: Culture**<br>- Tryptic soy broth (TSB) [E]<br>- Sheep blood [E, D]<br>- Chocolate [E]<br>- MacConkey [D]<br><br>**Environmental**<br>**Method: Culture**<br>- MacConkey agar [D]<br>- Xylose lysine deoxycholate (XLD) agar [S, D]<br>- Cystine lactose electrolyte deficient agar (CLED) [D]<br>- Sheep blood agar [E, D] | **Clinical and Environmental**<br>**Methodology: Presumptive phenotypic and biochemical identification**<br>- Fermentation method<br>- Urea 40% broth [D]<br>- Simmons citrate agar [S, D]<br>- Sulfide indole motility medium [D]<br>- Triple sugar iron agar [D]<br>- Klinger iron agar [D]<br>- Oxidase test reagents |

| | | | | | | |
|---|---|---|---|---|---|---|
| | | | | | Chocolate agar [E] | |
| 39 | **Clinical**<br>- Tracheal aspirates<br><br>**Environmental**<br>- Wastewater (hospital) | Clinical: 10<br>Environmental: 10 | Croatia | *Acinetobacter baumannii* | **Clinical**<br>- Previously isolated<br><br>**Environmental**<br>**Methodology: Membrane filtration and culture**<br>- CHROMagar Acinetobacter supplemented [S] with CR102 and cefsulodin sodium salt hydrate<br>Nutrient agar (NA) [E] | **Clinical**<br>**Methodology: Genotypic fingerprint**<br>- Previously identified<br>- PFGE<br><br>**Environmental**<br>**Methodology: Phenotypic and genotypic fingerprint**<br>- MALDI TOF MS<br>PFGE |
| 40 | **Clinical**<br>- Catheters, tracheobronchial, gastric, rectal, urine<br><br>**Environmental**<br>- Tap water | Clinical: 139<br>Environmental: 19 | Besançon, France | *Pseudomonas aeruginosa* | **Clinical and environmental**<br>**Methodology:** It was not specified | **Clinical and Environmental**<br>**Methodology: Genotypic identification**<br>PFGE |
| 41 | **Clinical**<br>- Nasal, ear, pus, wound, skin<br><br>**Environmental**<br>- Water samples: tap, borehole, tank, reservoir, well water | Clinical: 120<br>Environmental: 45 | Nigeria | *Staphylococcus aureus* | **Clinical**<br>**Methodology: Culture**<br>- Blood agar [E]<br>- Mannitol salt agar (MSA) [S, D]<br><br>**Environmental**<br>**Methodology: Culture**<br>Mannitol salt agar (MSA) [S, D] | **Clinical and Environmental**<br>Only the culture selective (presumptive phenotypic identification on selective and differential culture) |
| 42 | **Clinical**<br>- They only say that samples were from Austrian hospitals<br><br>**Environmental**<br>- River water | Clinical: 4<br>Environmental: 5 | Australia | *Klebsiella pneumoniae* | **Clinical**<br>It was not specified<br><br>**Environmental**<br>**Methodology: membrane filtration and culture**<br>- BBL fluid thioglycolate medium [E]<br>- Chromogenic media [S, D]<br>ChromID ESBL [S, D] | **Clinical**<br>- It was not specified<br><br>**Environmental**<br>**Methodology: Phenotypic and genotypic fingerprint**<br><br>- MALDI TOF MS<br>- WGS |
| 43 | **Clinical**<br>- Diarrheal stool | Clinical: 15<br>Environmental: 49 | India | *Klebsiella pneumoniae* | **Clinical**<br>**Methodology: Direct plating**<br>- Nutrient agar (NA) [E] | **Clinical and environmental:**<br>**Methodology: Presumptive phenotypic identification** |

| | | | | | | |
|---|---|---|---|---|---|---|
| | **Environmental**<br>- Well water (residential, market, hospital)<br>- Effluent (Fish market) | | | | - MacConkey (MA) [D]<br><br>**Environmental**<br>**Methodology: Direct plating**<br>- Tryptic soy agar (TSA) [E]<br>- MacConkey (MA) [D]<br>Nutrient agar (NA) [E] | Selective and differential medium. |
| 44 | **Clinical**<br>- Feces from healthy individuals and farmed animals<br>- Fish samples<br>- Pork and poultry meat from supermarkets<br><br>**Environmental**<br>- River and laker waters | Clinical: 108<br>Environmental: 11 | China | *Enterococcus (*including *Enterococcus faecium)* | **Clinical**<br>**Methodology: Culture**<br>- Luria-Bertani (LB) broth [E]<br>- Columbia agar base supplemented with sheep blood and florfenicol [S]<br>- Gram-staining<br><br>**Environmental**<br>**Methodology: Membrane filtration and culture**<br>- Luria-Bertani (LB) broth [E]<br>Columbia agar base supplemented with sheep blood and florfenicol [S] | **Clinical and environmental**<br>**Methodology: Phenotypic and Genotypic identification**<br>- MALDI-TOF MS<br>- WGS |
| 45 | **Clinical**<br>- Patients hospitalized<br><br>**Environmental**<br>- Raw hospital wastewater, inflow and outflow from hospital<br>- River water | Clinical: 110<br>Environmental: 231 | Czech Republi c | *Klebsiella* spp. | **Clinical**<br>**Methodology: Culture**<br>- MacConkey agar [S] supplemented with cefotaxime or meropenem.<br>**Environmental**<br>**Methodology: Membrane filtration and culture**<br>- Klebsiella ChromoSelect [S]<br>MacConkey [S] supplemented with cefotaxime or meropenem | **Clinical and Environmental**<br>**Methodology: Phenotypic and Genotypic identification**<br>- MALDI-TOF<br>WGS |
