## Supplementary material for "Antimicrobial resistance prevalence in clinical and aquatic environmental ESKAPE: a systematic review with meta-analysis": . Pooled antimicrobial resistance and heterogeneity metrics by site and dataset type

**Supplementary table 4**. Pooled antimicrobial resistance and heterogeneity metrics by site (clinical and environmental) and dataset type (all, with effluent, without effluent.

| Statistics | All datasets | | | Datasets with effluents | | | Datasets without effluents | | |
|---|---|---|---|---|---|---|---|---|---|
| | All dataset | Clinical | Environmental | All datasets | Clinical | Environmental | All datasets | Clinical | Environmental |
| $\tau^2$ | 14.11 | 9.62 | 17.24 | 11.58 | 12.17 | 10.66 | 19.15 | 6.19 | 43.87 |
| $I^2$ | 98.76 | 97.93 | 99.08 | 98.78 | 98.05 | 99.03 | 98.66 | 97.28 | 98.81 |
| $H^2$ | 80.45 | 48.32 | 108.54 | 81.77 | 51.41 | 102.88 | 74.68 | 36.78 | 84.07 |
| Wald | 4109.4 * | 1337.2* | 2706.6* | 3178.9 * | 674.21 * | 2476.2 * | 913.9* | 661.53* | 230.7* |
| LRT | 16888.6* | 5160.2* | 11375.8* | 12118.9 * | 3285.9 * | 8715.2 * | 4762.8* | 1857.8* | 2606.0* |
| Pooled mean | 0.46 | 0.67 | 0.24 | 0.42 | 0.59 | 0.28 | 0.51 | 0.76 | 0.15 |
| LCI | 0.36 | 0.55 | 0.14 | 0.30 | 0.58 | 0.17 | 0.36 | 0.63 | 0.03 |
| HCI | 0.57 | 0.77 | 0.39 | 0.54 | 0.59 | 0.44 | 0.69 | 0.85 | 0.49 |

* *P* value < 0.001. The articles that considered effluents in the environmental samples were: 23,25,26,30,32,34,38,40. The studies that not included effluent in the environmental samples were: 24,25,27,29,33,35–37,39
