## Supplementary material for "Antimicrobial resistance prevalence in clinical and aquatic environmental ESKAPE: a systematic review with meta-analysis": Publication heterogeneity on the patterns of antimicrobial resistance in clinical and environmental samples

**Supplementary table 5.** Publication heterogeneity on the patterns of antimicrobial resistance in clinical and environmental samples

| Antibiotic class | Site | N | Egger´s regression test | | Funnel plot |
|---|---|---|---|---|---|
| | | | t value | *p* value | |
| Aminoglycoside | Clinical | 12 | -0.97 | 0.34 | Symmetrical |
| Aminoglycoside | Environmental | 12 | -2.60 | **0.02** | **Asymmetrical** |
| Amphenicol | Clinical | 6 | 0.85 | 0.44 | Symmetrical |
| Amphenicol | Environmental | 6 | 0.45 | 0.67 | Symmetrical |
| Beta-lactam | Clinical | 16 | 4.53 | **< 0.001** | **Asymmetrical** |
| Beta-lactam | Environmental | 16 | 1.35 | 0.18 | Symmetrical |
| Beta-lactam/BLI | Clinical | 7 | 0.45 | 0.66 | Symmetrical |
| Beta-lactam/BLI | Environmental | 8 | 0.34 | 0.74 | Symmetrical |
| Fluoroquinolone | Clinical | 13 | 0.86 | 0.40 | Symmetrical |
| Fluoroquinolone | Environmental | 13 | -3.41 | **< 0.001** | **Asymmetrical** |
| Macrolide | Clinical | 5 | -0.43 | 0.69 | Symmetrical |
| Macrolide | Environmental | 5 | 2.74 | 0.07 | Symmetrical |
| Sulfonamide | Clinical | 6 | 1.20 | 0.28 | Symmetrical |
| Sulfonamide | Environmental | 6 | -0.68 | 0.53 | Symmetrical |
| Tetracycline | Clinical | 7 | 0.52 | 0.63 | Symmetrical |
| Tetracycline | Environmental | 8 | -0.87 | 0.42 | Symmetrical |

N = Number of studies in meta-analysis
