## Supplementary material for "Antimicrobial resistance prevalence in clinical and aquatic environmental ESKAPE: a systematic review with meta-analysis": Comparison of pooled antimicrobial resistance proportions between clinical and environmental bacterial isolates by antimicrobial class and agent

| | | | | | | | | | |
|---|---|---|---|---|---|---|---|---|---|
| **Beta Lactams**<br>Clinical: 74.1%<br>Environmental: 46.6% | | **Cephalosporins 1a** | **Cephalosporins 2a** | **Cephalosporins 3a** | | | | | **Cephalosporins 4a** |
| | | **Cefazolin** | **Cefuroxime** | **Ceftazidime** | **Cefotaxime** | **Ceftriaxone** | **Cefpodoxime-proxetil** | **Cefoperazone** | **Cefepime** |
| | **Clinical** | 88.9 | 93.9 | 77.9 | 90.4 | 59.7 | 34.8 | 99.5 | 81.2 |
| | **Environmental** | 99.2 | 95.4 | 39.2 | 78.9 | 41.7 | 99.3 | 97.8 | 27.5 |
| | | **Penicillin** | | | | | **Monobactams** | | |
| | | **Ampicillin** | **Cloxacillin** | **Penicillin** | **Piperacillin** | **Ticarcillin** | **Aztreonam** | | |
| | **Clinical** | 70.9 | 99.6 | 20.1 | 73.3 | 83.5 | 90.7 | | |
| | **Environmental** | 52.1 | 48 | 22.1 | 25.4 | 49.6 | 30.5 | | |
| | | **Carbapenems** | | | | | | | |
| | | **Ertapenem** | **Imipenem** | **Meropenem** | | | | | |
| | **Clinical** | 37.2 | 45.4 | 41.5 | | | | | |
| | **Environmental** | 27.6 | 11.3 | 15.5 | | | | | |
| **Beta-Lactams/ BLI**<br><br>Clinical: 64.1%<br>Environmental: 51.6% | | **Penicillin** | | | | | | | |
| | | **Ampicillin/ sulbactam** | **Amoxicillin/ Clavulanic acid** | **Ticarcillin/ clavulanic acid** | **Amoxicillin/ clavulanic acid** | **Piperacillin/ tazobactam** | | | |
| | **Clinical** | 73.4 | 83.1 | 97.6 | 91.7 | 34.8 | | | |
| | **Environmental** | 92.2 | 82.2 | 66.7 | 48 | 12.8 | | | |
| | | **Cephalosporins 3a** | | | | | | | |
| | | **Cefoperazone/ sulbactam** | **Ceftazidime/ clavulanic acid** | | | | | | |
| | **Clinical** | 60.0 | 66.7 | | | | | | |
| | **Environmental** | 71.4 | 24.5 | | | | | | |
| **Aminoglycoside**<br>Clinical: 42.2%<br>Environmental: 13.5% | | **Amikacin** | **Gentamicin** | **Netilmicin** | **Streptomycin** | **Tobramycin** | | | |
| | **Clinical** | 8.9 | 50.3 | 42.7 | 65.6 | 45.1 | | | |
| | **Environmental** | 5.2 | 16.9 | 10.3 | 26.6 | 24.8 | | | |
| **Fluoroquinolone**<br>Clinical: 60.9%<br>Environmental: 17.8% | | **Ciprofloxacin** | **Levofloxacin** | **Norfloxacin** | **Ofloxacin** | | | | |
| | **Clinical** | 61.3 | 83.7 | 2.4 | 63.1 | | | | |
| | **Environmental** | 34.2 | 22.5 | 0.6 | 3.1 | | | | |
| **Amphenicol**<br>Clinical: 48.8%<br>Environmental: 28.1% | | **Chloramphenicol** | | | | | | | |
| | **Clinical** | 48.8 | | | | | | | |
| | **Environmental** | 28.1 | | | | | | | |
| **Glycopeptide**<br>Clinical: 12.3%<br>Environmental: 1.3% | | **Teicoplanin** | **Vancomycin** | | | | | | |
| | **Clinical** | 18.7 | 9.2 | | | | | | |
| | **Environmental** | 1.4 | 1.3 | | | | | | |
| **Glycylcyclines**<br>Clinical: 46.9%<br>Environmental: 35.4% | | **Tigecycline** | | | | | | | |
| | **Clinical** | 46.9 | | | | | | | |
| | **Environmental** | 35.4 | | | | | | | |
| **Sulfonamide**<br>Clinical: 82.7%<br>Environmental: 51.2% | | **Trimethoprim / sulfametoxazole** | | | | | | | |
| | **Clinical** | 82.7 | | | | | | | |
| | **Environmental** | 51.2 | | | | | | | |
| **Macrolide**<br>Clinical: 90.8%<br>Environmental: 86.4% | | **Erythromycin** | | | | | | | |
| | **Clinical** | 90.8 | | | | | | | |
| | **Environmental** | 86.4 | | | | | | | |
| **Nitrofuran** | | **Nitrofurantoin** | | | | | | | |

| | | |
|---|---|---|
| Clinical: 7.5%<br>Environmental: 14.5% | **Clinical** | 7.5 |
| | **Environmental** | 14.5 |
| **Oxazolidinone**<br>Clinical: 11.8%<br>Environmental: 11.2% | | **Linezolid** |
| | **Clinical** | 11.8 |
| | **Environmental** | 11.2 |
| **Polymyxin**<br>Clinical: 3.3%<br>Environmental: 9.5% | | **Colistin** |
| | **Clinical** | 3.3 |
| | **Environmental** | 9.5 |
| **Rifamycin**<br>Clinical: 3.3%<br>Environmental: 34.3% | | **Rifampin** |
| | **Clinical** | 3.3 |
| | **Environmental** | 34.3 |
| **Streptogramin**<br>Clinical: 3.4%<br>Environmental: 3.4% | | **Quinupristin/ Dalfopristin** |
| | **Clinical** | 2.1 |
| | **Environmental** | 3.4 |
| **Tetracycline**<br>Clinical: 71.4%<br>Environmental: 44.8% | | **Tetracycline** |
| | **Clinical** | 71.4 |
| | **Environmental** | 44.8 |
