## Supplementary material for "Antimicrobial resistance prevalence in clinical and aquatic environmental ESKAPE: a systematic review with meta-analysis": Forest plots of pooled antimicrobial resistance proportions by antibiotic class comparing clinical and environmental bacterial isolates

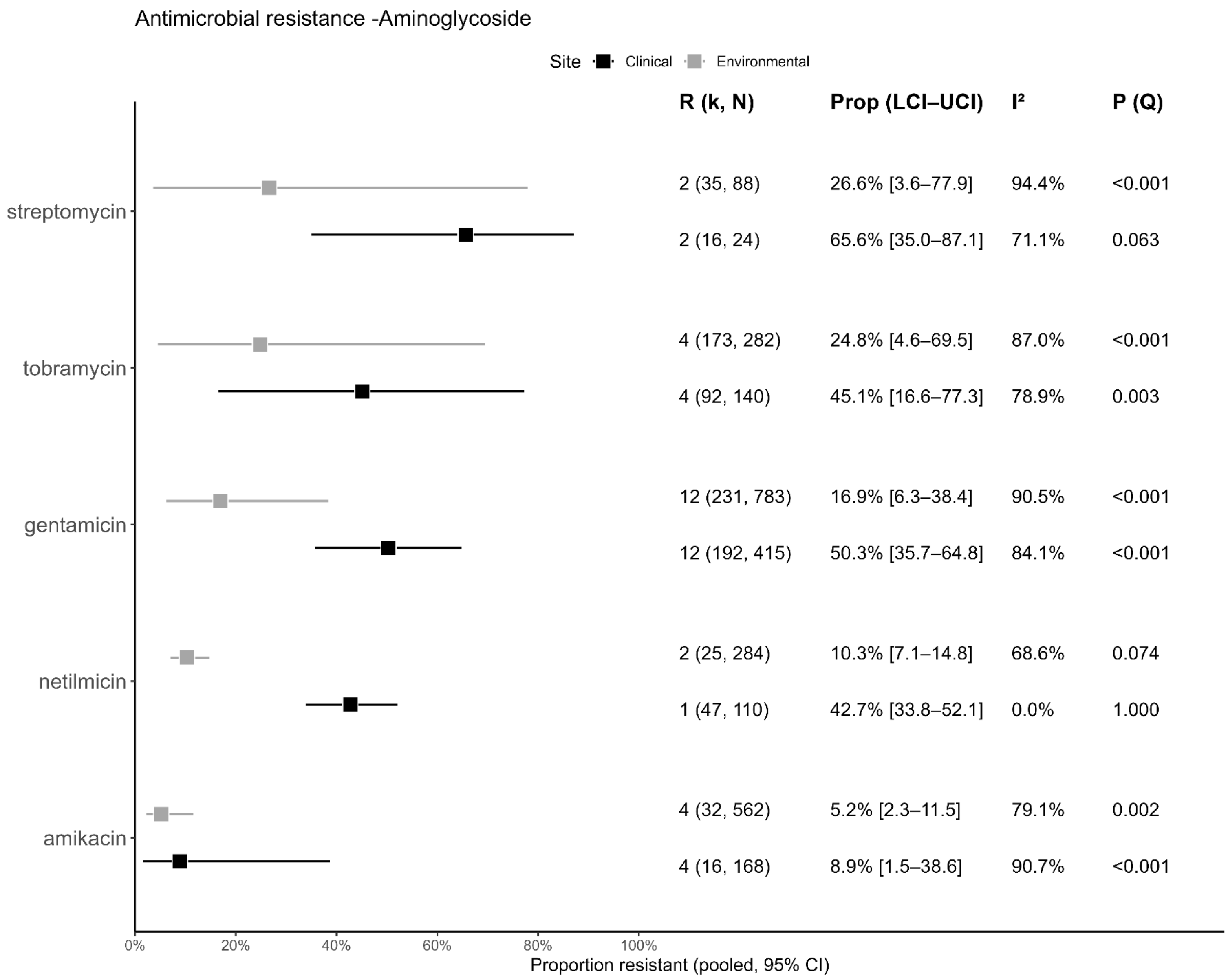

Antimicrobial resistance -Aminoglycoside
Site
Clinical
Environmental
R (k, N)
Prop (LCI–UCI)
I²
P (Q)
streptomycin
2 (35, 88) 26.6% [3.6–77.9] 94.4% <0.001
2 (16, 24) 65.6% [35.0–87.1] 71.1% 0.063
tobramycin
4 (173, 282) 24.8% [4.6–69.5] 87.0% <0.001
4 (92, 140) 45.1% [16.6–77.3] 78.9% 0.003
gentamicin
12 (231, 783) 16.9% [6.3–38.4] 90.5% <0.001
12 (192, 415) 50.3% [35.7–64.8] 84.1% <0.001
netilmicin
2 (25, 284) 10.3% [7.1–14.8] 68.6% 0.074
1 (47, 110) 42.7% [33.8–52.1] 0.0% 1.000
amikacin
4 (32, 562) 5.2% [2.3–11.5] 79.1% 0.002
4 (16, 168) 8.9% [1.5–38.6] 90.7% <0.001
0%
20%
40%
60%
80%
100%
Proportion resistant (pooled, 95% CI)

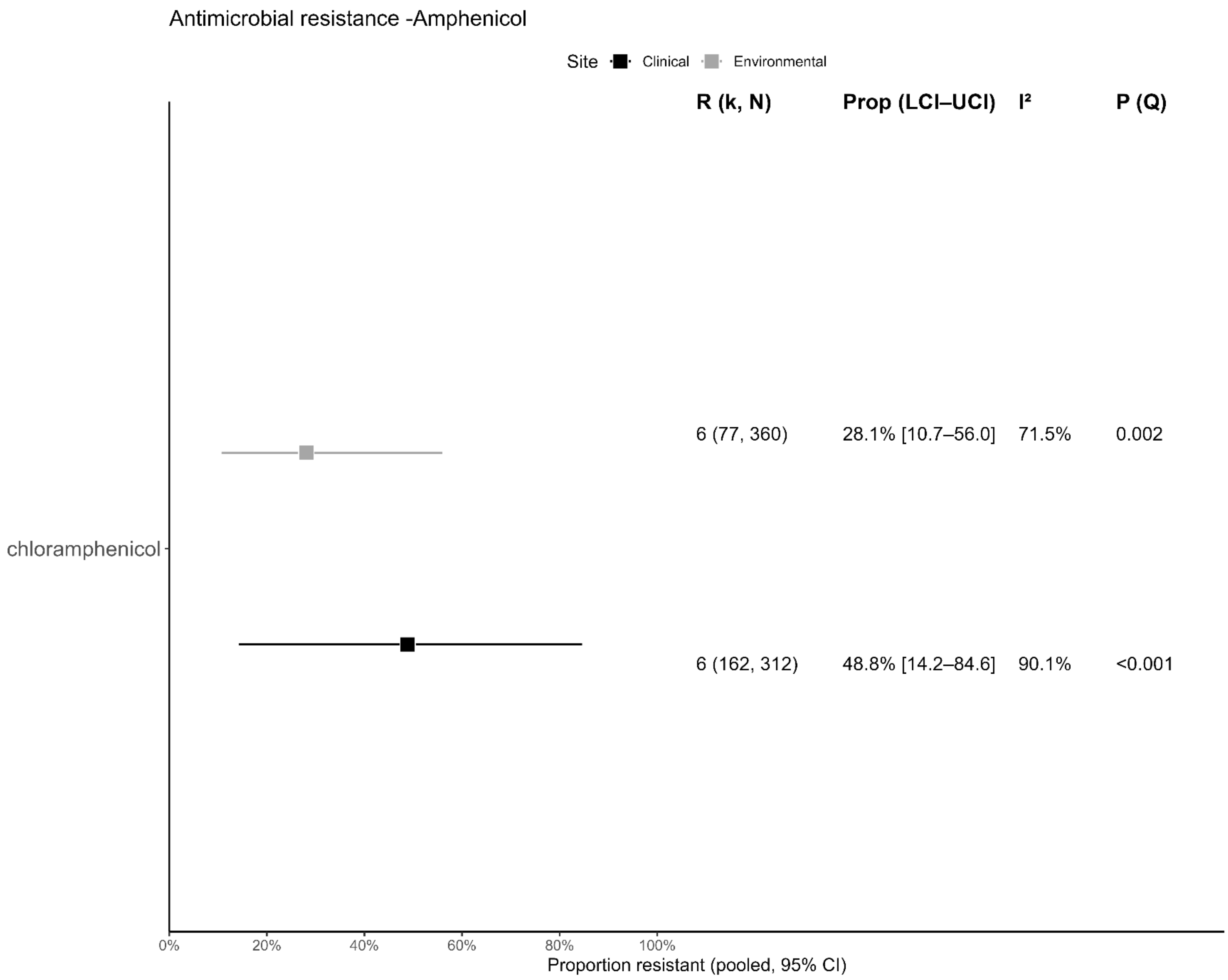
Antimicrobial resistance -Amphenicol
Site Clinical Environmental
R (k, N)
Prop (LCI–UCI)
I²
P (Q)
6 (77, 360)
28.1% [10.7–56.0]
71.5%
0.002
chloramphenicol
6 (162, 312)
48.8% [14.2–84.6]
90.1%
<0.001
0%
20%
40%
60%
80%
100%
Proportion resistant (pooled, 95% CI)

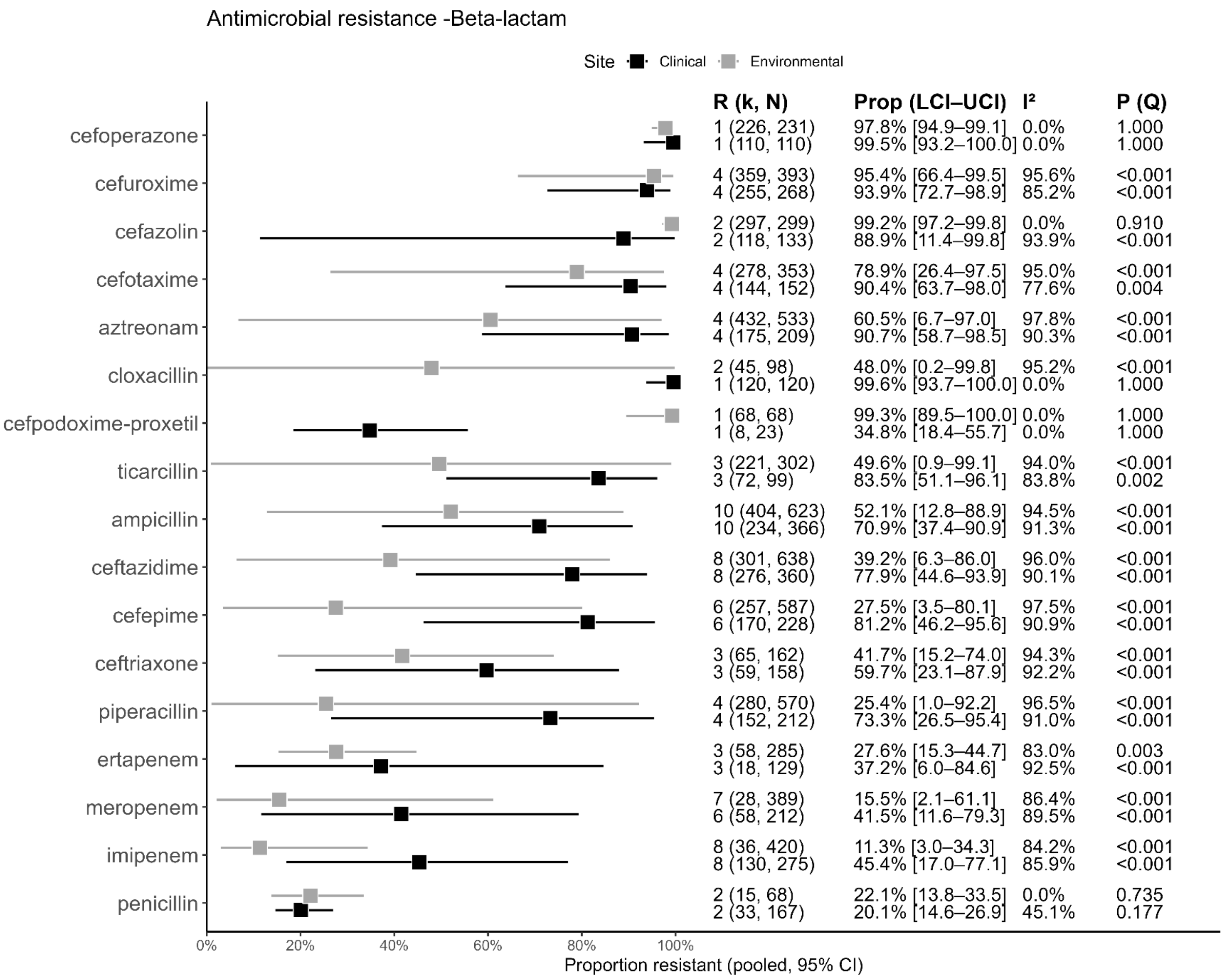
Antimicrobial resistance -Beta-lactam
Site Clinical Environmental
R (k, N) Prop (LCI–UCI) I² P (Q)
cefoperazone
1 (226, 231) 97.8% [94.9–99.1] 0.0% 1.000
1 (110, 110) 99.5% [93.2–100.0] 0.0% 1.000
cefuroxime
4 (359, 393) 95.4% [66.4–99.5] 95.6% <0.001
4 (255, 268) 93.9% [72.7–98.9] 85.2% <0.001
cefazolin
2 (297, 299) 99.2% [97.2–99.8] 0.0% 0.910
2 (118, 133) 88.9% [11.4–99.8] 93.9% <0.001
cefotaxime
4 (278, 353) 78.9% [26.4–97.5] 95.0% <0.001
4 (144, 152) 90.4% [63.7–98.0] 77.6% 0.004
aztreonam
4 (432, 533) 60.5% [6.7–97.0] 97.8% <0.001
4 (175, 209) 90.7% [58.7–98.5] 90.3% <0.001
cloxacillin
2 (45, 98) 48.0% [0.2–99.8] 95.2% <0.001
1 (120, 120) 99.6% [93.7–100.0] 0.0% 1.000
cefpodoxime-proxetil
1 (68, 68) 99.3% [89.5–100.0] 0.0% 1.000
1 (8, 23) 34.8% [18.4–55.7] 0.0% 1.000
ticarcillin
3 (221, 302) 49.6% [0.9–99.1] 94.0% <0.001
3 (72, 99) 83.5% [51.1–96.1] 83.8% 0.002
ampicillin
10 (404, 623) 52.1% [12.8–88.9] 94.5% <0.001
10 (234, 366) 70.9% [37.4–90.9] 91.3% <0.001
ceftazidime
8 (301, 638) 39.2% [6.3–86.0] 96.0% <0.001
8 (276, 360) 77.9% [44.6–93.9] 90.1% <0.001
cefepime
6 (257, 587) 27.5% [3.5–80.1] 97.5% <0.001
6 (170, 228) 81.2% [46.2–95.6] 90.9% <0.001
ceftriaxone
3 (65, 162) 41.7% [15.2–74.0] 94.3% <0.001
3 (59, 158) 59.7% [23.1–87.9] 92.2% <0.001
piperacillin
4 (280, 570) 25.4% [1.0–92.2] 96.5% <0.001
4 (152, 212) 73.3% [26.5–95.4] 91.0% <0.001
ertapenem
3 (58, 285) 27.6% [15.3–44.7] 83.0% 0.003
3 (18, 129) 37.2% [6.0–84.6] 92.5% <0.001
meropenem
7 (28, 389) 15.5% [2.1–61.1] 86.4% <0.001
6 (58, 212) 41.5% [11.6–79.3] 89.5% <0.001
imipenem
8 (36, 420) 11.3% [3.0–34.3] 84.2% <0.001
8 (130, 275) 45.4% [17.0–77.1] 85.9% <0.001
penicillin
2 (15, 68) 22.1% [13.8–33.5] 0.0% 0.735
2 (33, 167) 20.1% [14.6–26.9] 45.1% 0.177
0% 20% 40% 60% 80% 100%
Proportion resistant (pooled, 95% CI)

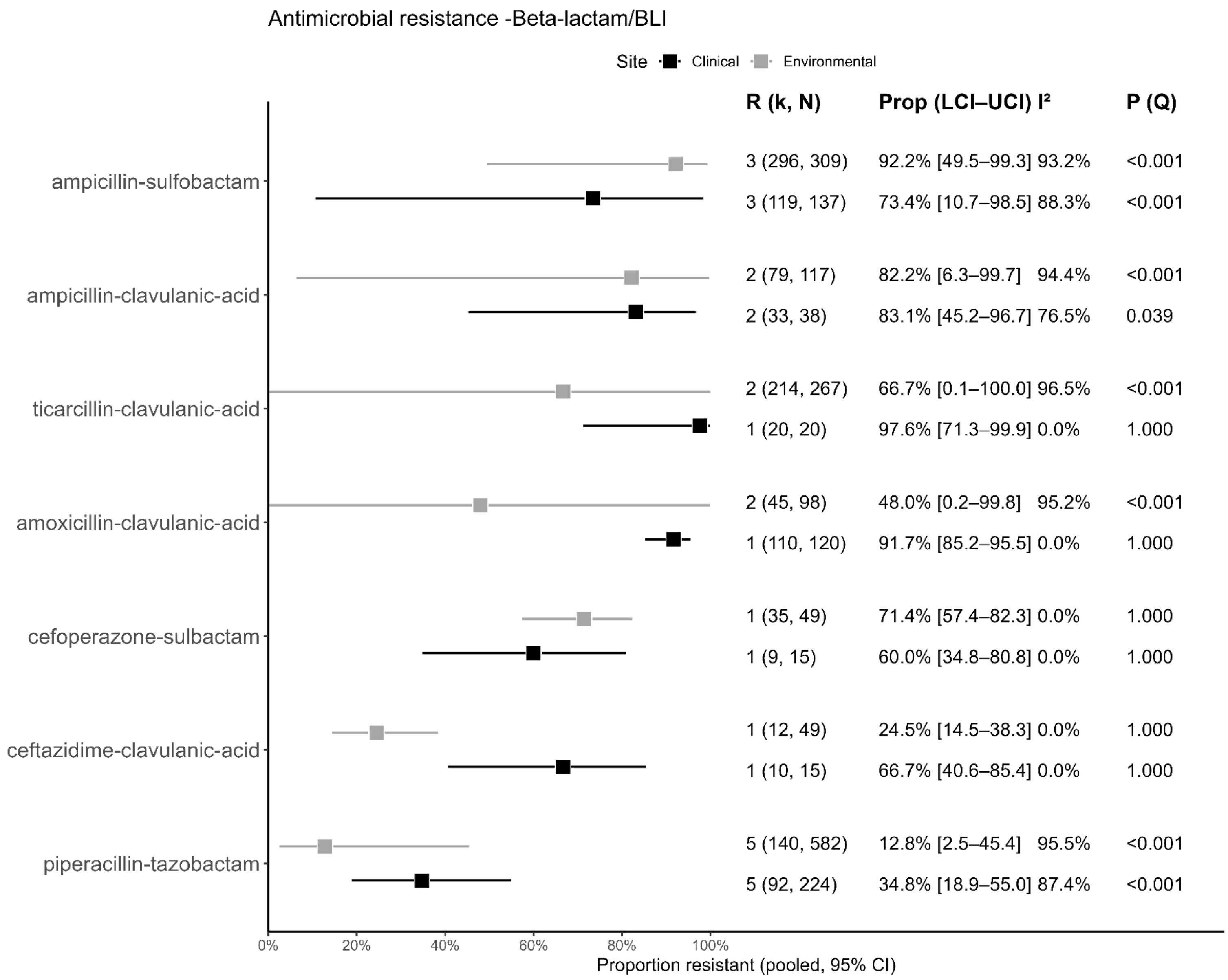

Antimicrobial resistance -Beta-lactam/BLI
Site Clinical Environmental
R (k, N)
Prop (LCI–UCI)
I²
P (Q)
ampicillin-sulfobactam
3 (296, 309) 92.2% [49.5–99.3] 93.2% <0.001
3 (119, 137) 73.4% [10.7–98.5] 88.3% <0.001
ampicillin-clavulanic-acid
2 (79, 117) 82.2% [6.3–99.7] 94.4% <0.001
2 (33, 38) 83.1% [45.2–96.7] 76.5% 0.039
ticarcillin-clavulanic-acid
2 (214, 267) 66.7% [0.1–100.0] 96.5% <0.001
1 (20, 20) 97.6% [71.3–99.9] 0.0% 1.000
amoxicillin-clavulanic-acid
2 (45, 98) 48.0% [0.2–99.8] 95.2% <0.001
1 (110, 120) 91.7% [85.2–95.5] 0.0% 1.000
cefoperazone-sulbactam
1 (35, 49) 71.4% [57.4–82.3] 0.0% 1.000
1 (9, 15) 60.0% [34.8–80.8] 0.0% 1.000
ceftazidime-clavulanic-acid
1 (12, 49) 24.5% [14.5–38.3] 0.0% 1.000
1 (10, 15) 66.7% [40.6–85.4] 0.0% 1.000
piperacillin-tazobactam
5 (140, 582) 12.8% [2.5–45.4] 95.5% <0.001
5 (92, 224) 34.8% [18.9–55.0] 87.4% <0.001
0% 20% 40% 60% 80% 100%
Proportion resistant (pooled, 95% CI)

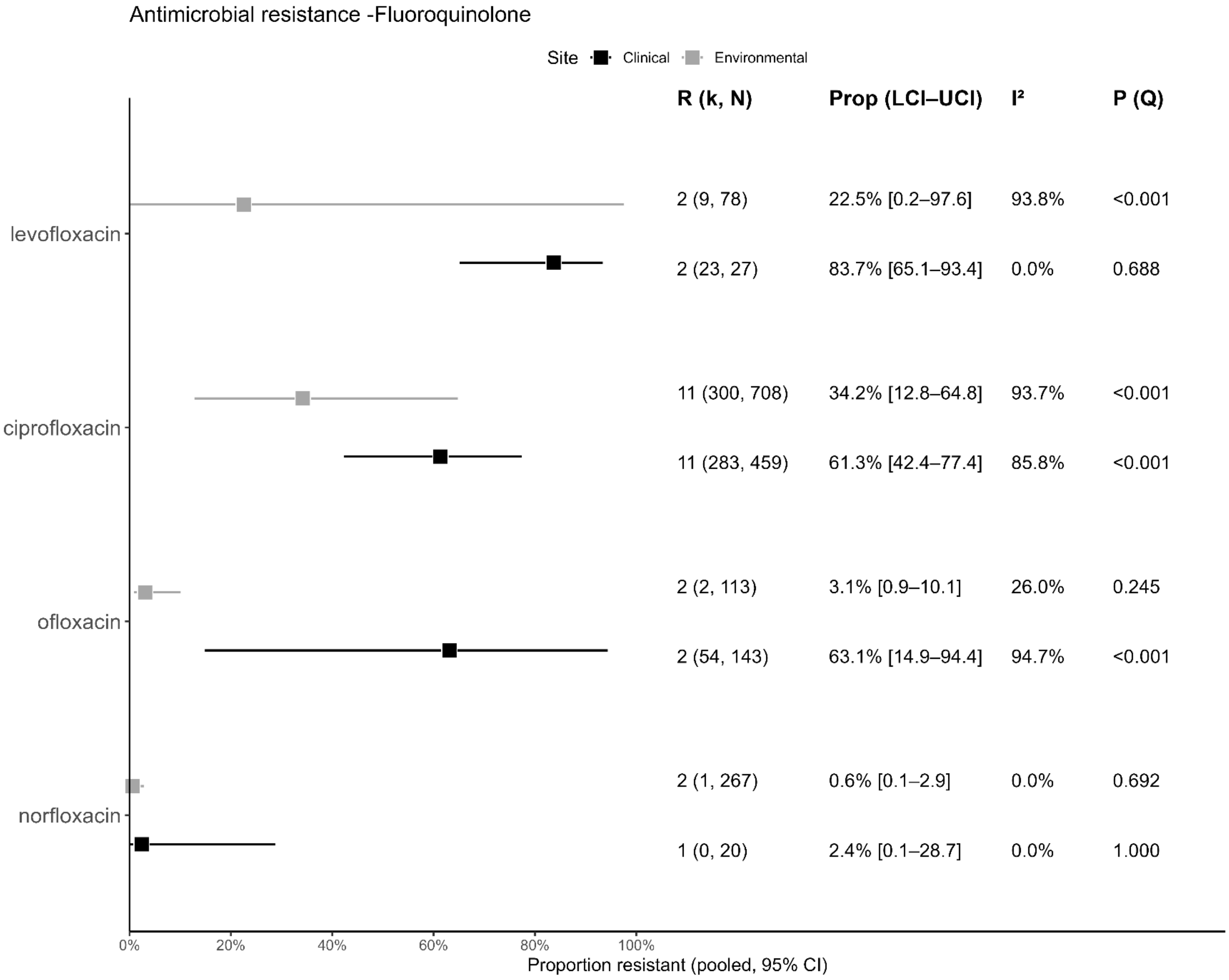
Antimicrobial resistance -Fluoroquinolone
Site Clinical Environmental
R (k, N)
Prop (LCI–UCI)
I²
P (Q)
levofloxacin
2 (9, 78) 22.5% [0.2–97.6] 93.8% <0.001
2 (23, 27) 83.7% [65.1–93.4] 0.0% 0.688
ciprofloxacin
11 (300, 708) 34.2% [12.8–64.8] 93.7% <0.001
11 (283, 459) 61.3% [42.4–77.4] 85.8% <0.001
ofloxacin
2 (2, 113) 3.1% [0.9–10.1] 26.0% 0.245
2 (54, 143) 63.1% [14.9–94.4] 94.7% <0.001
norfloxacin
2 (1, 267) 0.6% [0.1–2.9] 0.0% 0.692
1 (0, 20) 2.4% [0.1–28.7] 0.0% 1.000
0%
20%
40%
60%
80%
100%
Proportion resistant (pooled, 95% CI)

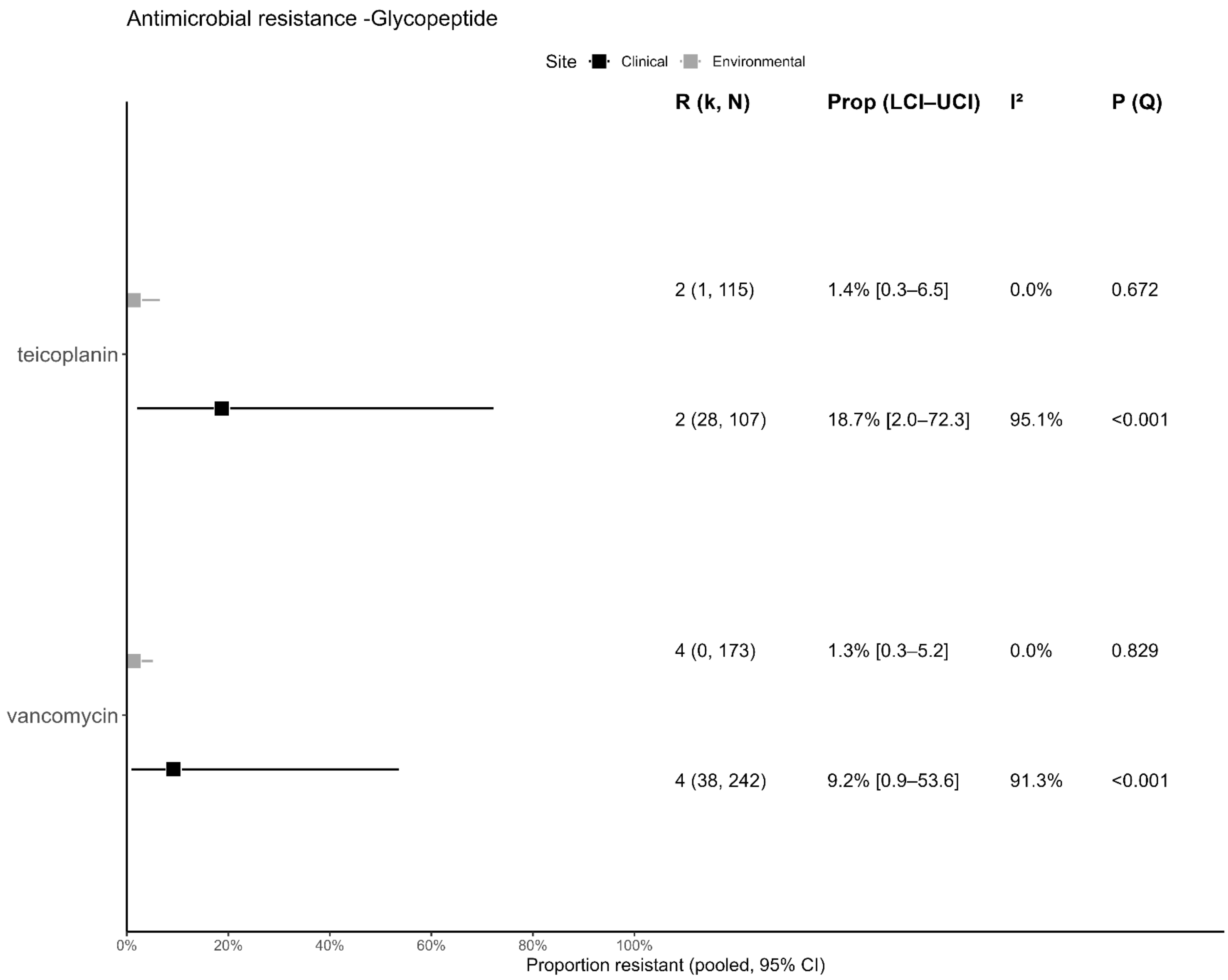
Antimicrobial resistance -Glycopeptide
Site
Clinical
Environmental
R (k, N)
Prop (LCI–UCI)
I²
P (Q)
teicoplanin
2 (1, 115)
1.4% [0.3–6.5]
0.0%
0.672
2 (28, 107)
18.7% [2.0–72.3]
95.1%
<0.001
vancomycin
4 (0, 173)
1.3% [0.3–5.2]
0.0%
0.829
4 (38, 242)
9.2% [0.9–53.6]
91.3%
<0.001
0%
20%
40%
60%
80%
100%
Proportion resistant (pooled, 95% CI)

Antimicrobial resistance -Glycylcycline

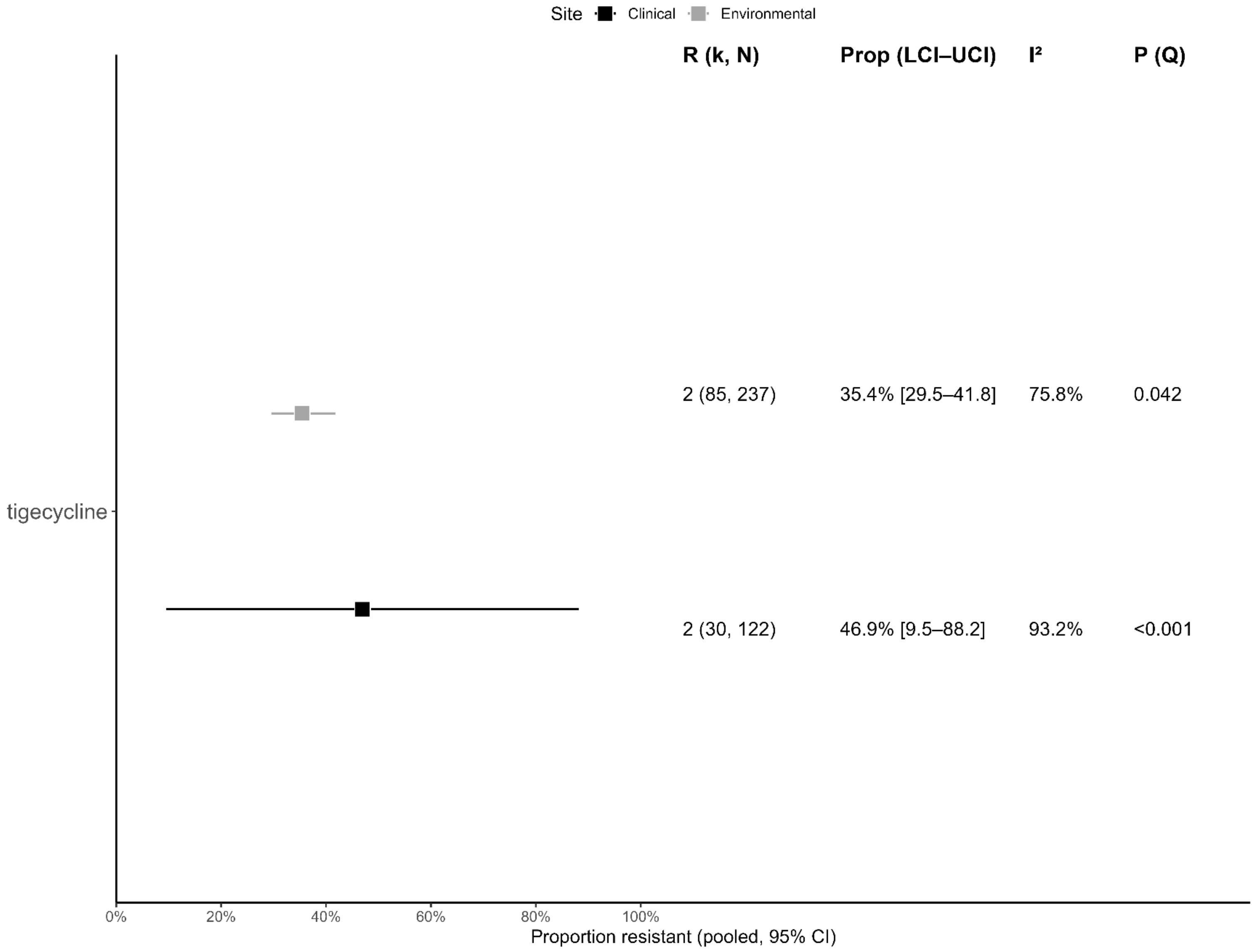

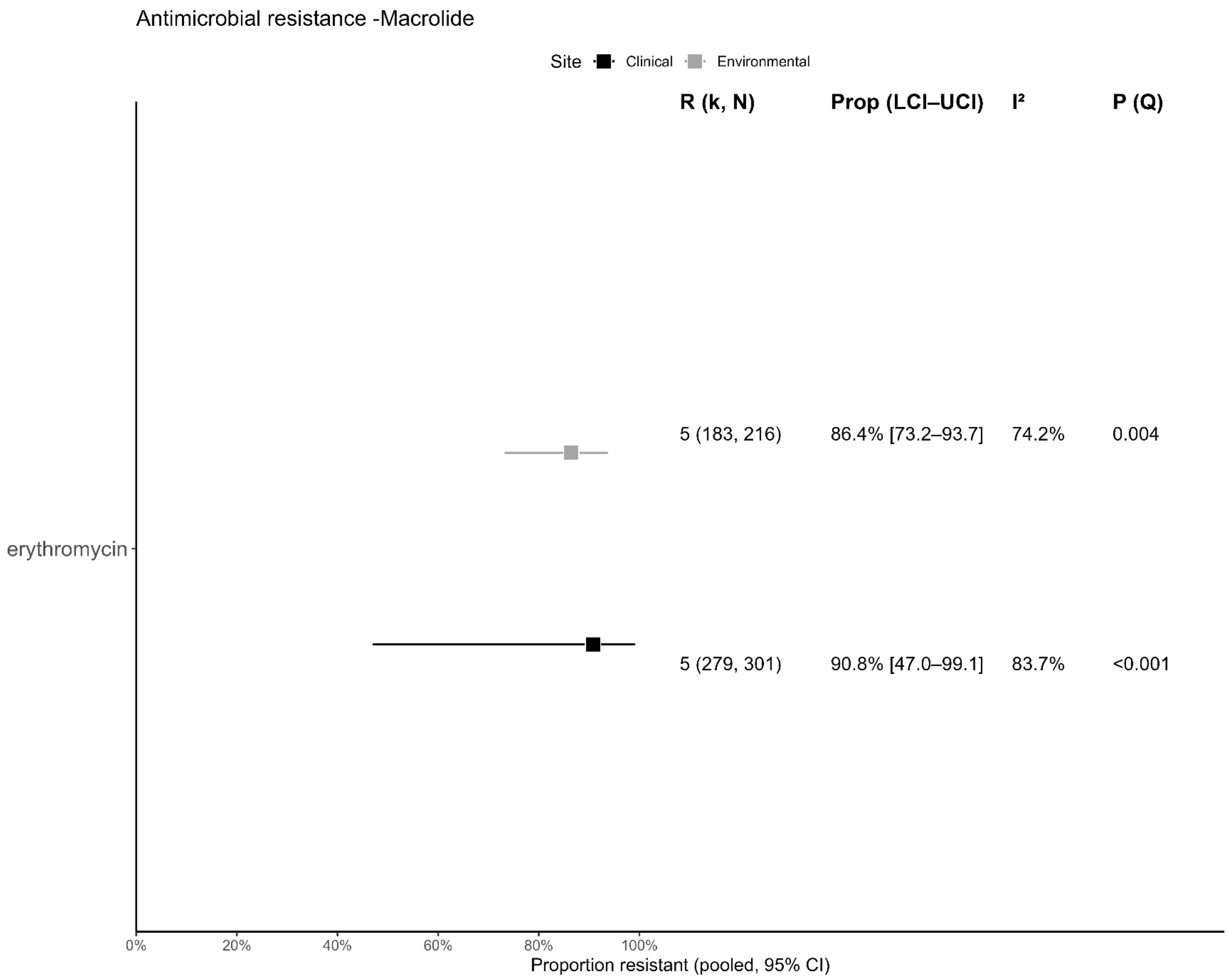
Antimicrobial resistance -Macrolide
Site
Clinical
Environmental
R (k, N)
Prop (LCI–UCI)
I²
P (Q)
5 (183, 216)
86.4% [73.2–93.7]
74.2%
0.004
erythromycin
5 (279, 301)
90.8% [47.0–99.1]
83.7%
<0.001
0%
20%
40%
60%
80%
100%
Proportion resistant (pooled, 95% CI)

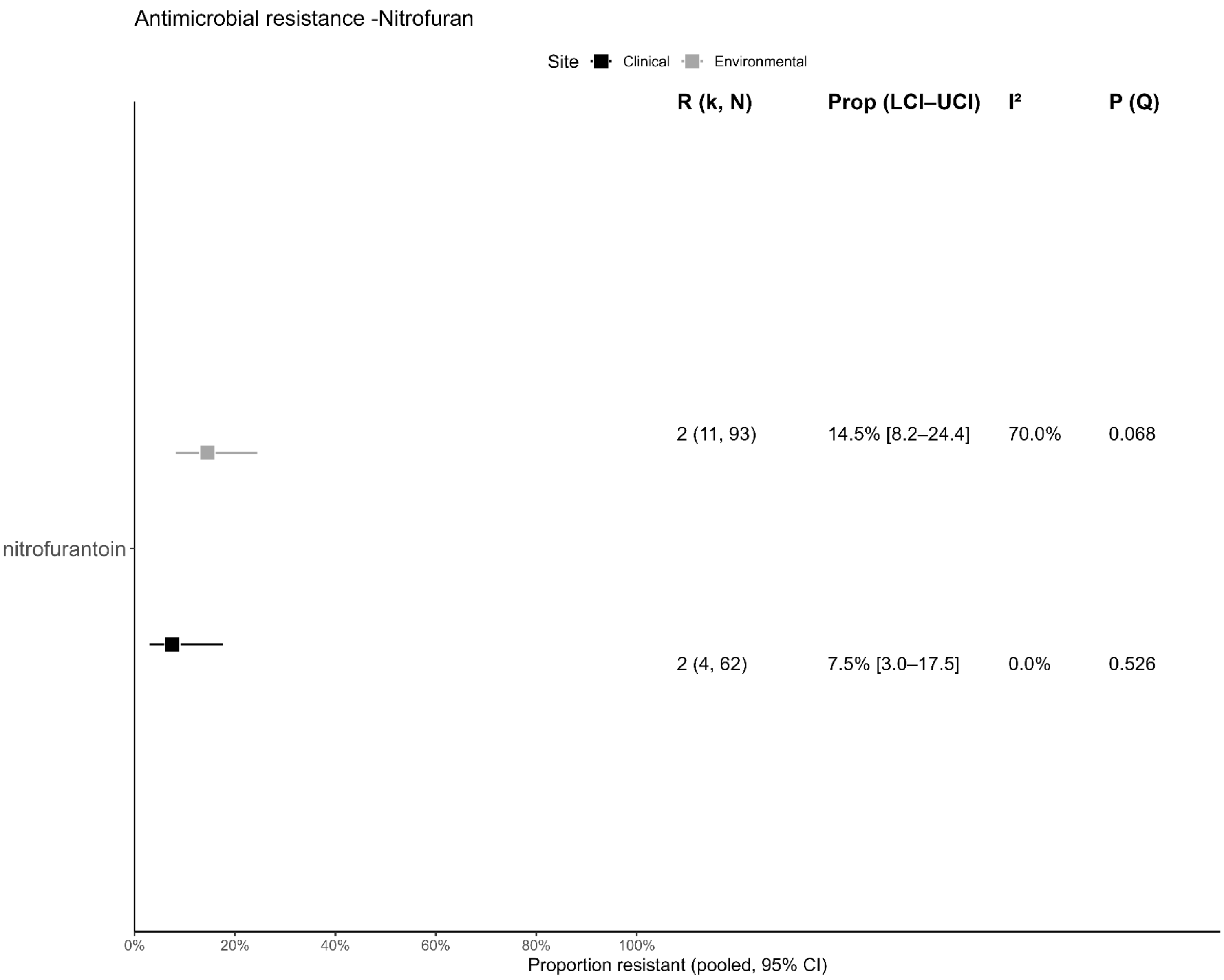
Antimicrobial resistance -Nitrofuran
Site
Clinical
Environmental
R (k, N)
Prop (LCI–UCI)
I²
P (Q)
2 (11, 93)
14.5% [8.2–24.4]
70.0%
0.068
nitrofurantoin
2 (4, 62)
7.5% [3.0–17.5]
0.0%
0.526
0%
20%
40%
60%
80%
100%
Proportion resistant (pooled, 95% CI)

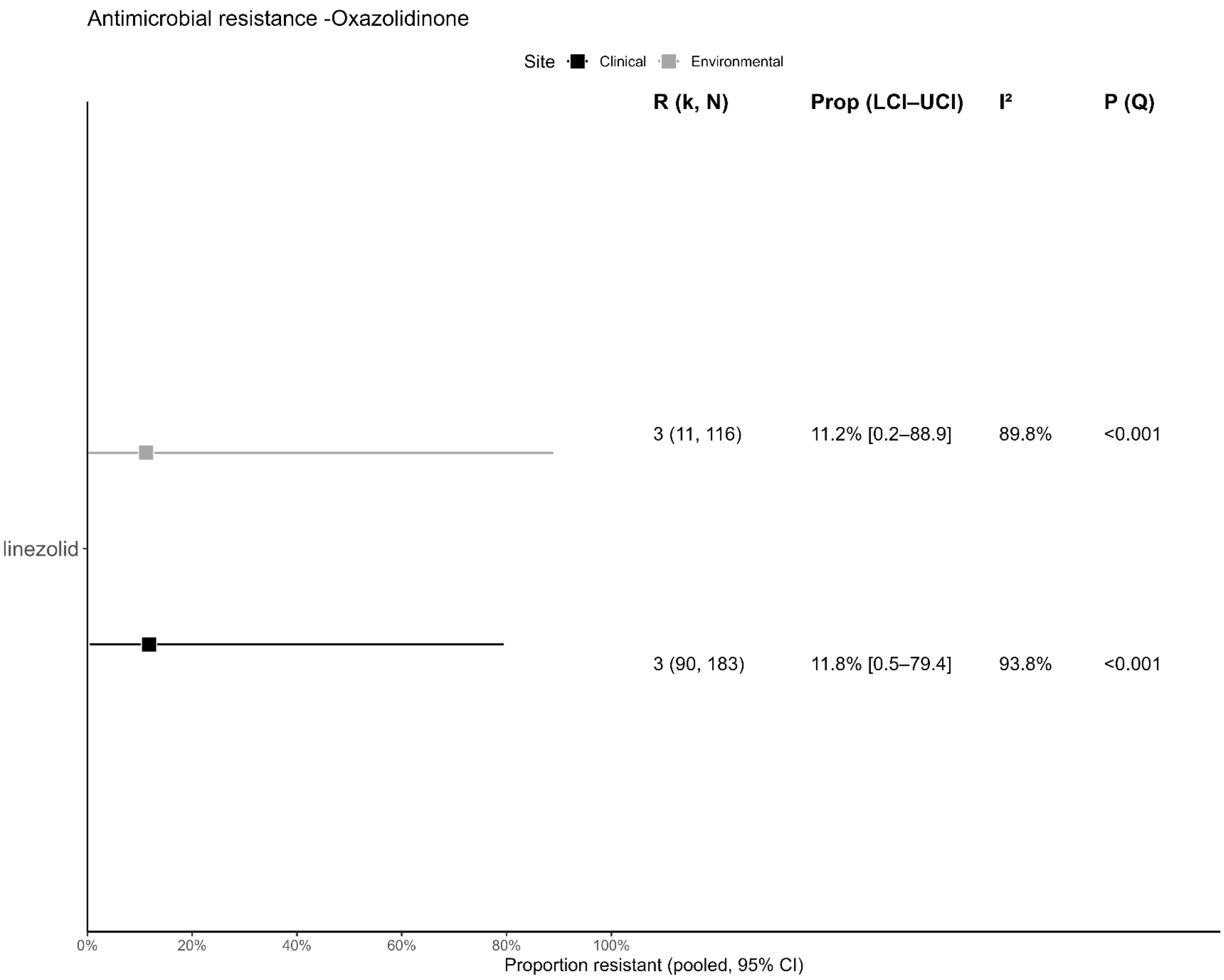
Antimicrobial resistance -Oxazolidinone
Site
Clinical
Environmental
R (k, N)
Prop (LCI–UCI)
I²
P (Q)
3 (11, 116)
11.2% [0.2–88.9]
89.8%
<0.001
linezolid
3 (90, 183)
11.8% [0.5–79.4]
93.8%
<0.001
0%
20%
40%
60%
80%
100%
Proportion resistant (pooled, 95% CI)

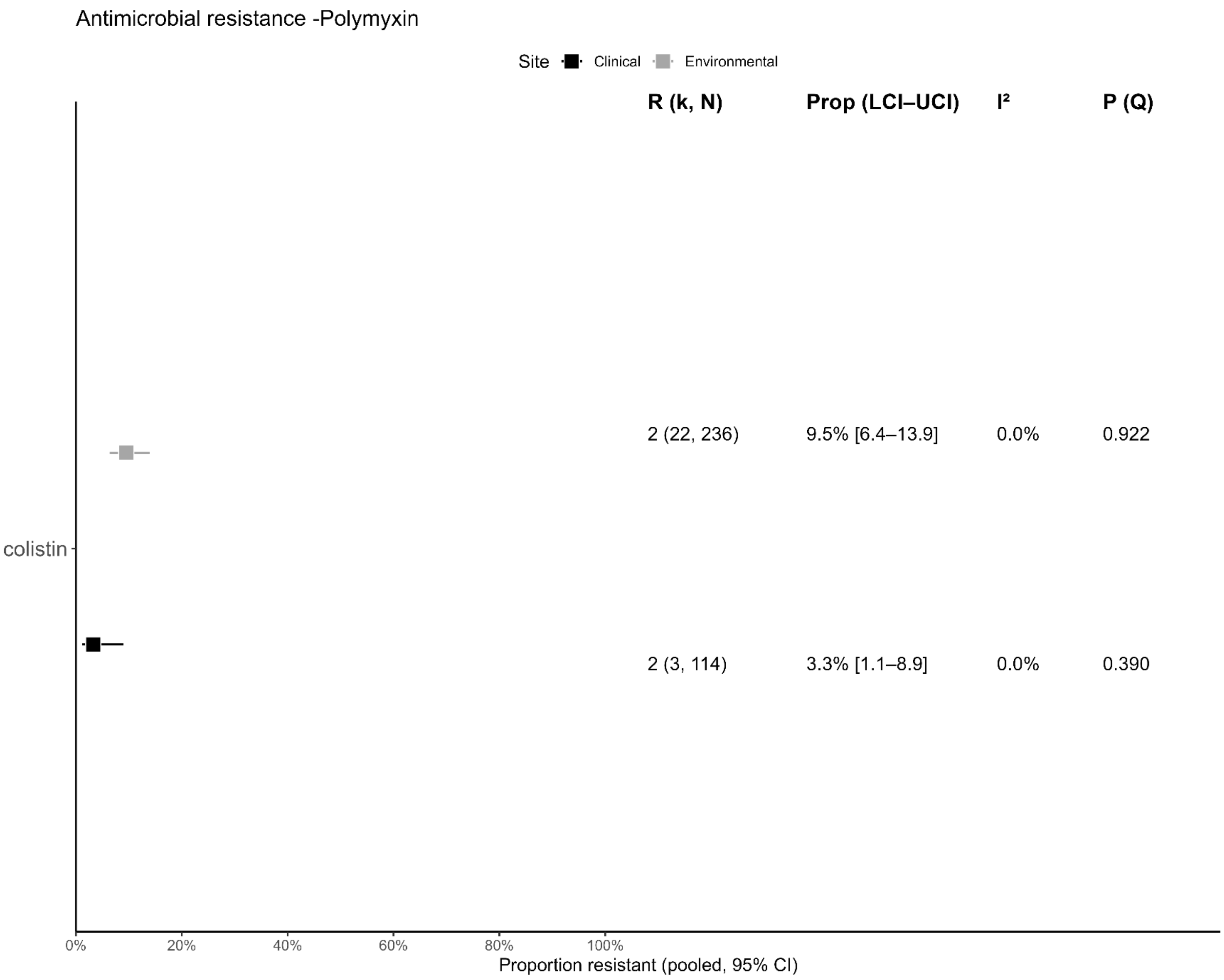
Antimicrobial resistance -Polymyxin
Site
Clinical
Environmental
R (k, N)
Prop (LCI–UCI)
I²
P (Q)
2 (22, 236)
9.5% [6.4–13.9]
0.0%
0.922
colistin
2 (3, 114)
3.3% [1.1–8.9]
0.0%
0.390
0%
20%
40%
60%
80%
100%
Proportion resistant (pooled, 95% CI)

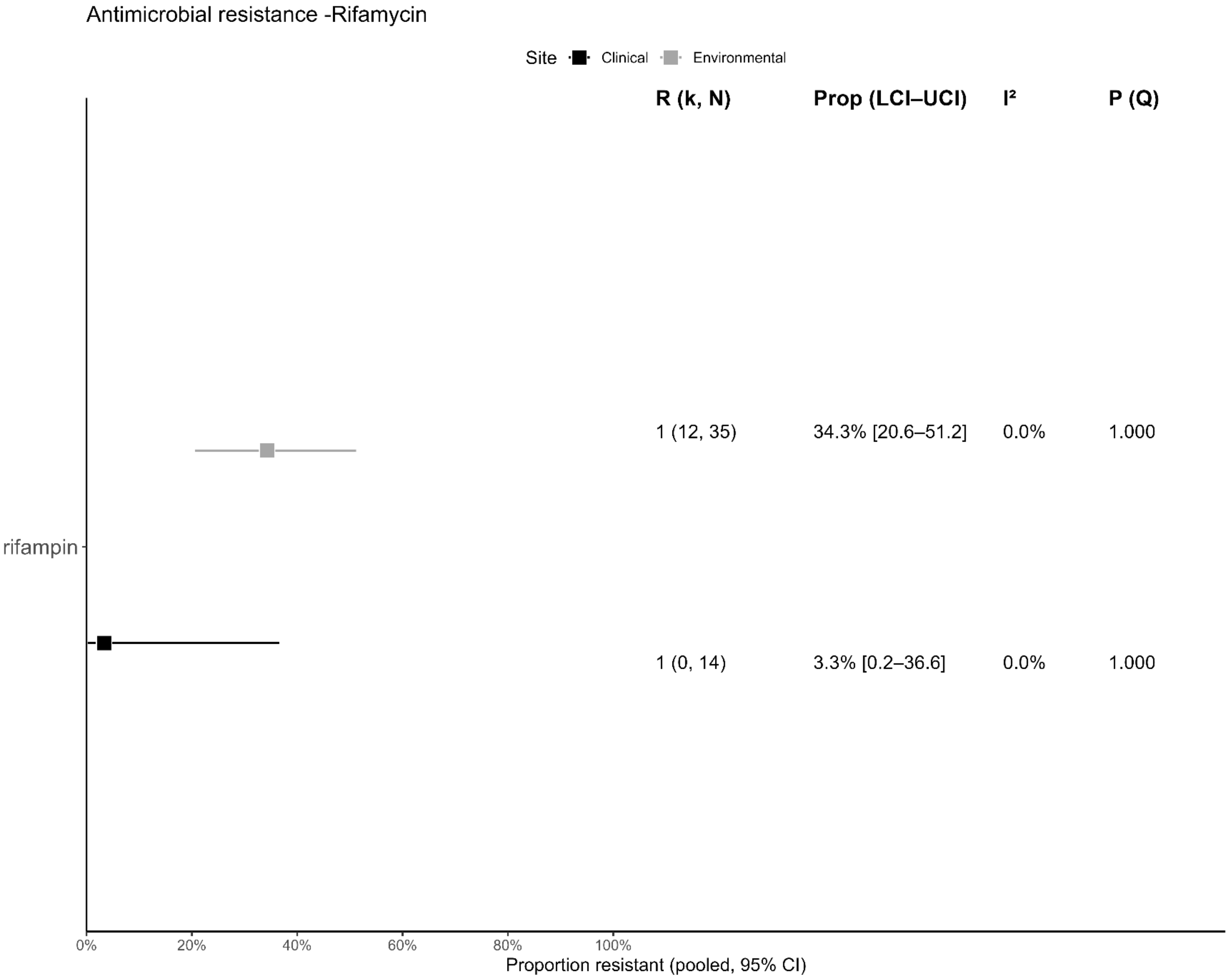
Antimicrobial resistance -Rifamycin
Site
Clinical
Environmental
R (k, N)
Prop (LCI–UCI)
I²
P (Q)
1 (12, 35)
34.3% [20.6–51.2]
0.0%
1.000
rifampin
1 (0, 14)
3.3% [0.2–36.6]
0.0%
1.000
0%
20%
40%
60%
80%
100%
Proportion resistant (pooled, 95% CI)

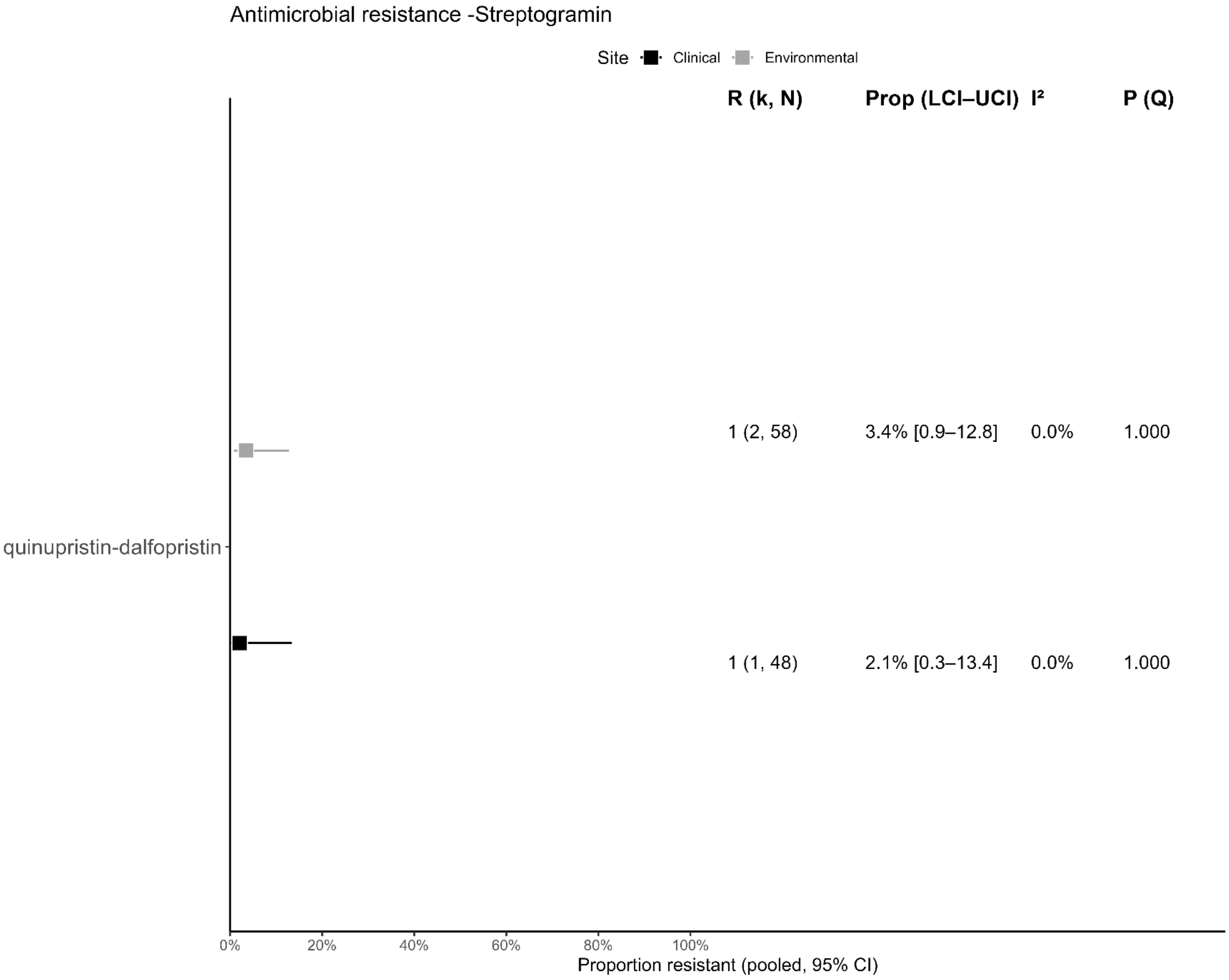

Antimicrobial resistance -Streptogramin
Site Clinical Environmental
R (k, N)
Prop (LCI–UCI)
I²
P (Q)
1 (2, 58)
3.4% [0.9–12.8]
0.0%
1.000
quinupristin-dalfopristin
1 (1, 48)
2.1% [0.3–13.4]
0.0%
1.000
0%
20%
40%
60%
80%
100%
Proportion resistant (pooled, 95% CI)

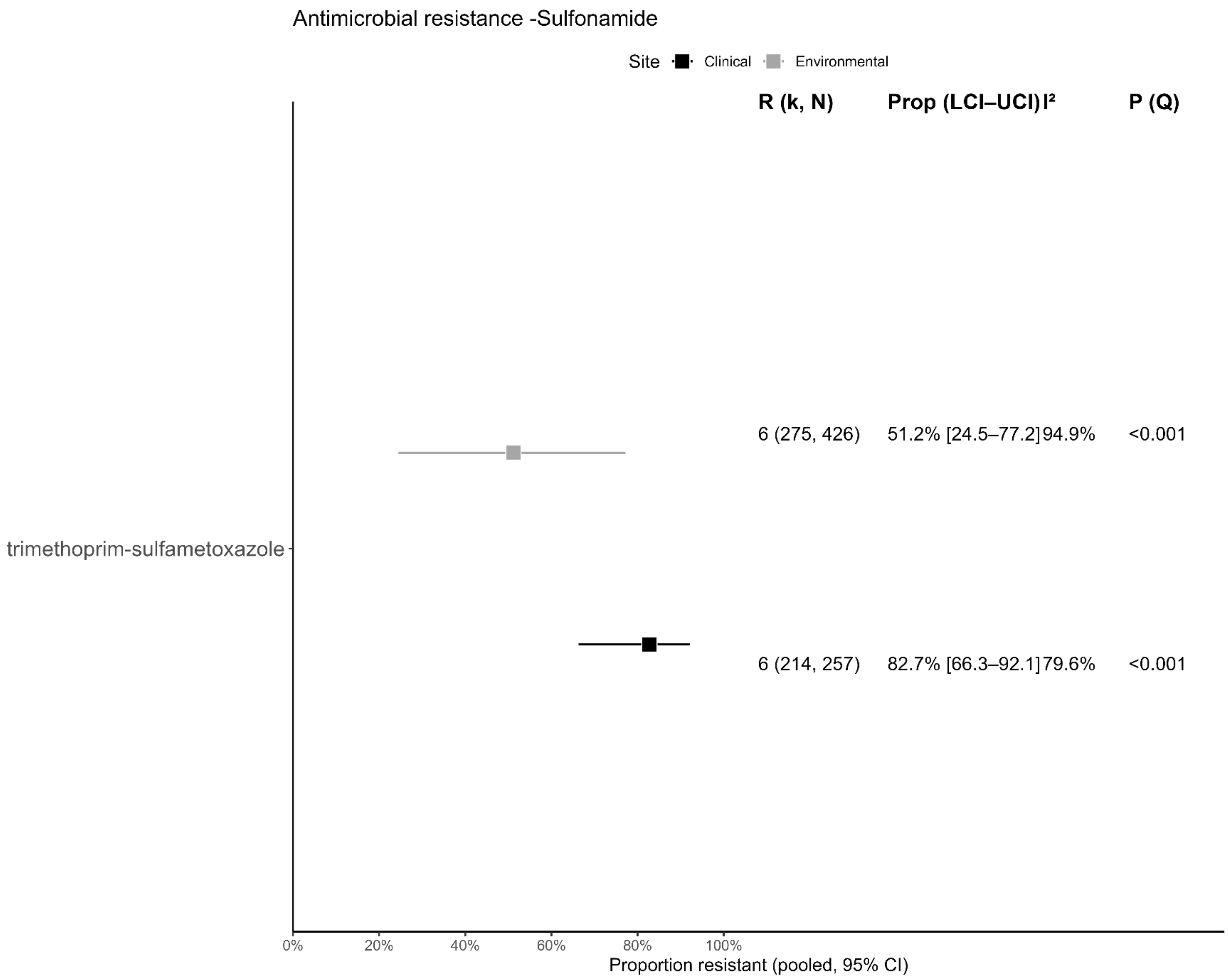
Antimicrobial resistance -Sulfonamide
Site
Clinical
Environmental
R (k, N)
Prop (LCI–UCI)
I²
P (Q)
6 (275, 426)
51.2% [24.5–77.2]
94.9%
<0.001
trimethoprim-sulfametoxazole
6 (214, 257)
82.7% [66.3–92.1]
79.6%
<0.001
0%
20%
40%
60%
80%
100%
Proportion resistant (pooled, 95% CI)

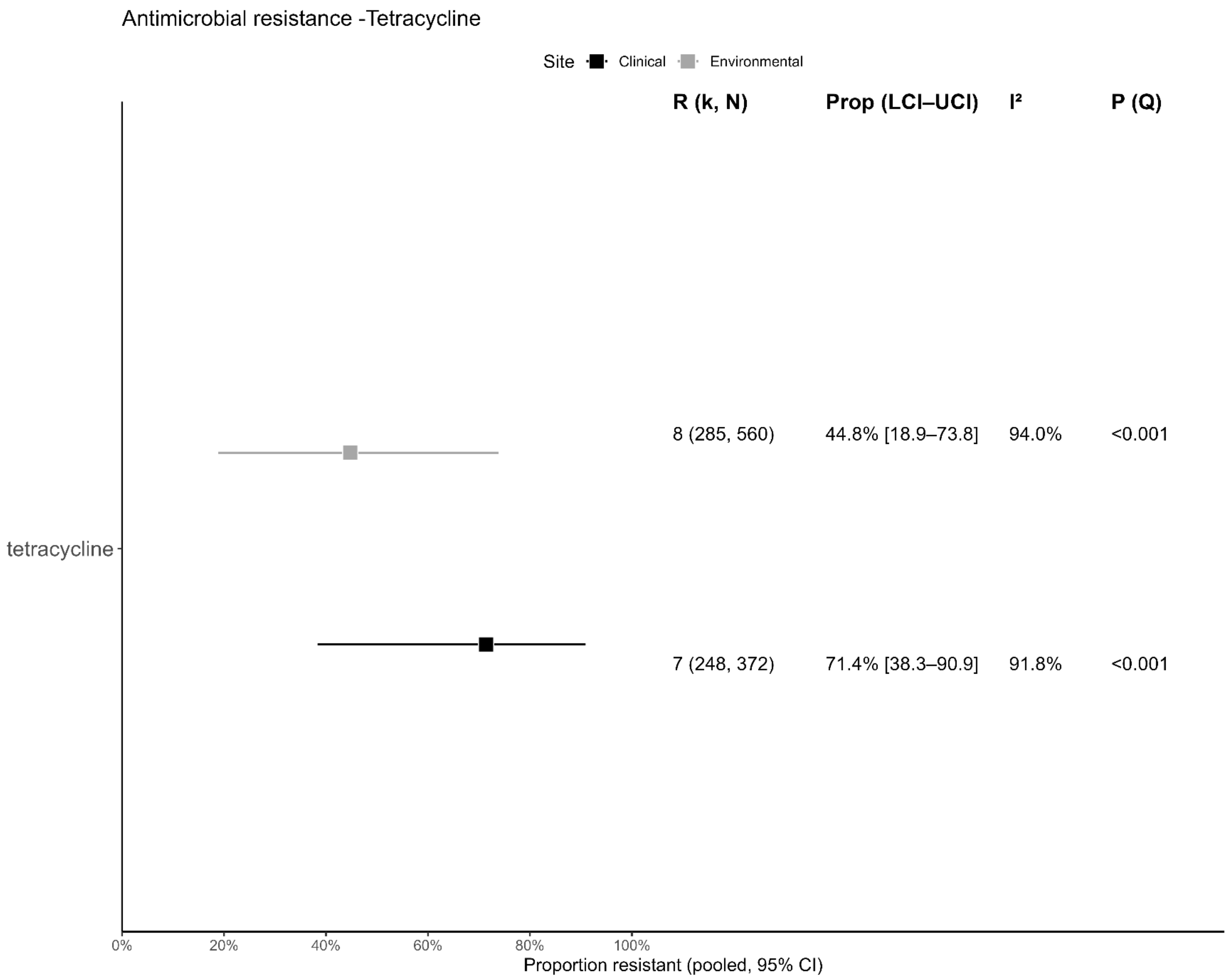

Antimicrobial resistance -Tetracycline
Site
Clinical
Environmental
R (k, N)
Prop (LCI–UCI)
I²
P (Q)
8 (285, 560)
44.8% [18.9–73.8]
94.0%
<0.001
tetracycline
7 (248, 372)
71.4% [38.3–90.9]
91.8%
<0.001
0%
20%
40%
60%
80%
100%
Proportion resistant (pooled, 95% CI)

**Supplementary Figure 1.** Forest plot of the proportion of antibiotic resistance by antibiotic.
