## Supplementary figures and images for "Antimicrobial resistance prevalence in clinical and aquatic environmental ESKAPE: a systematic review with meta-analysis"

### Funnel plots of pooled antimicrobial resistance estimates for clinical and environmental isolates by antibiotic class

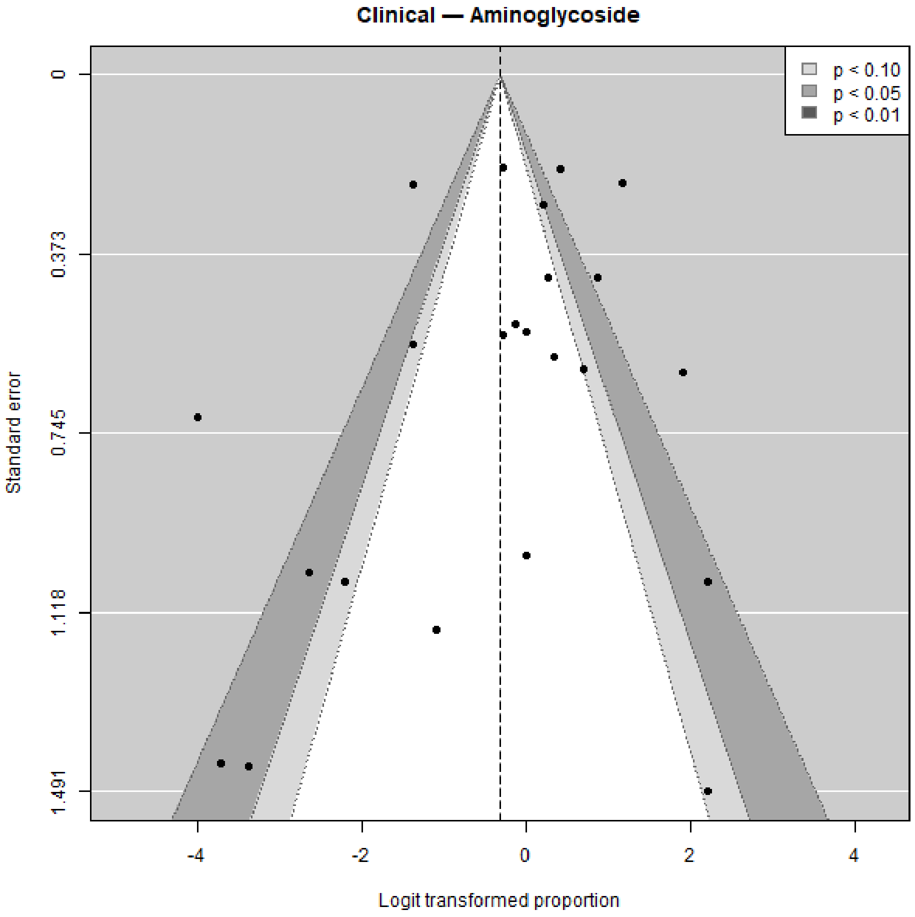

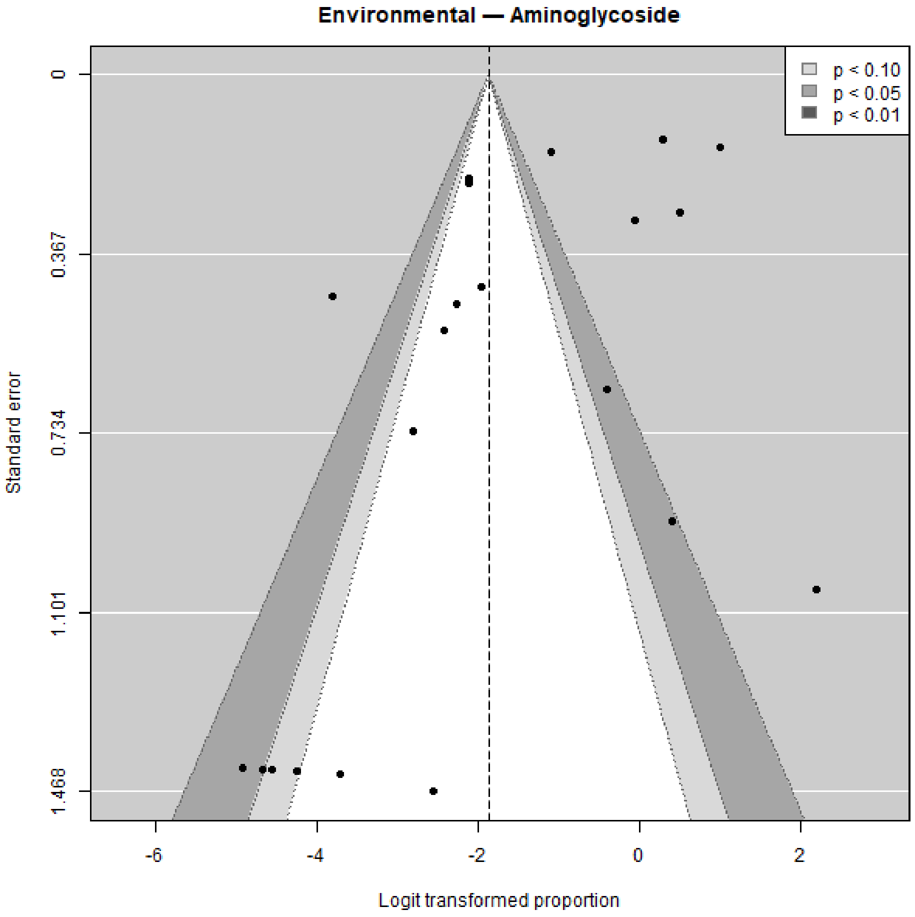

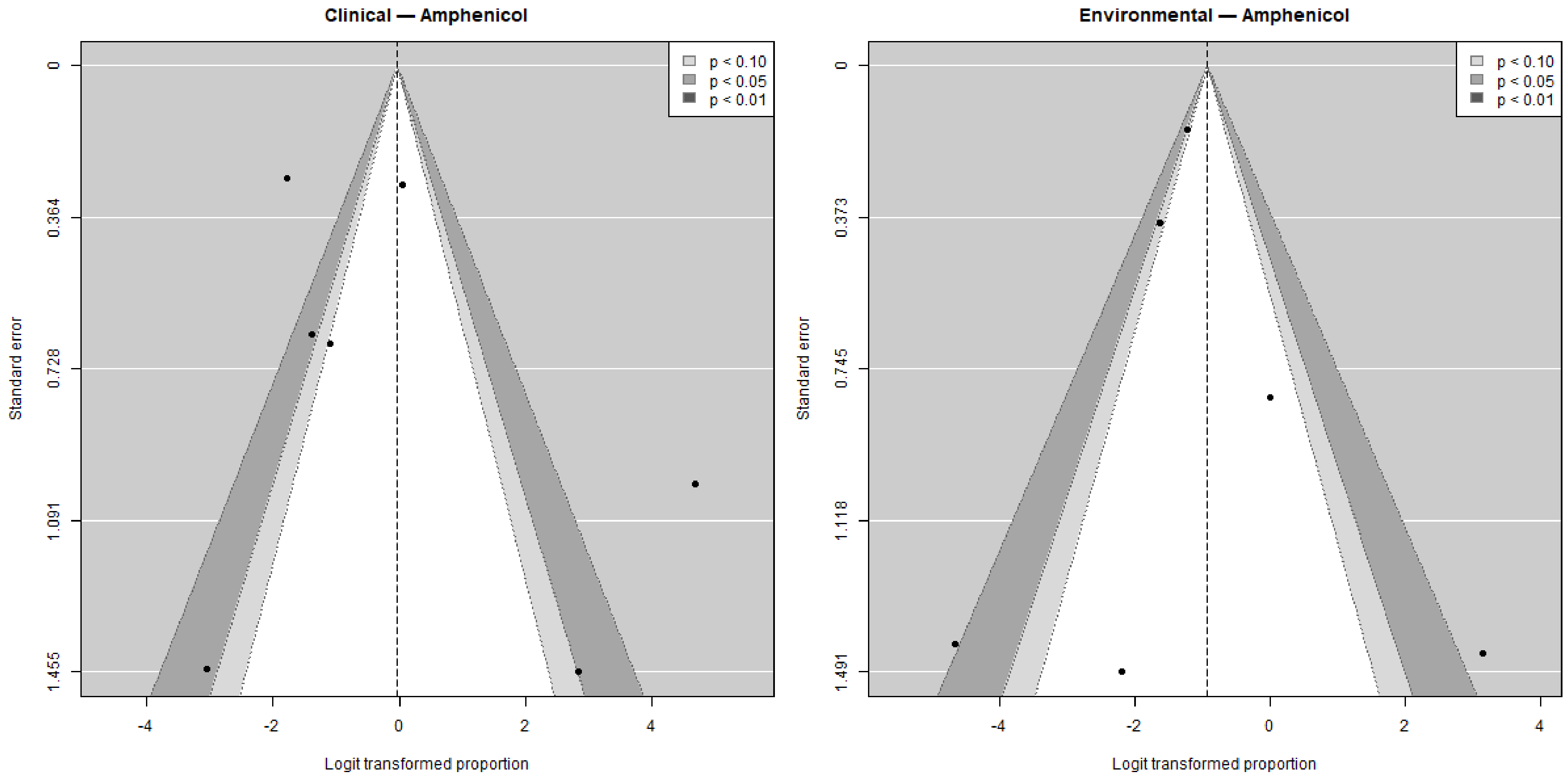

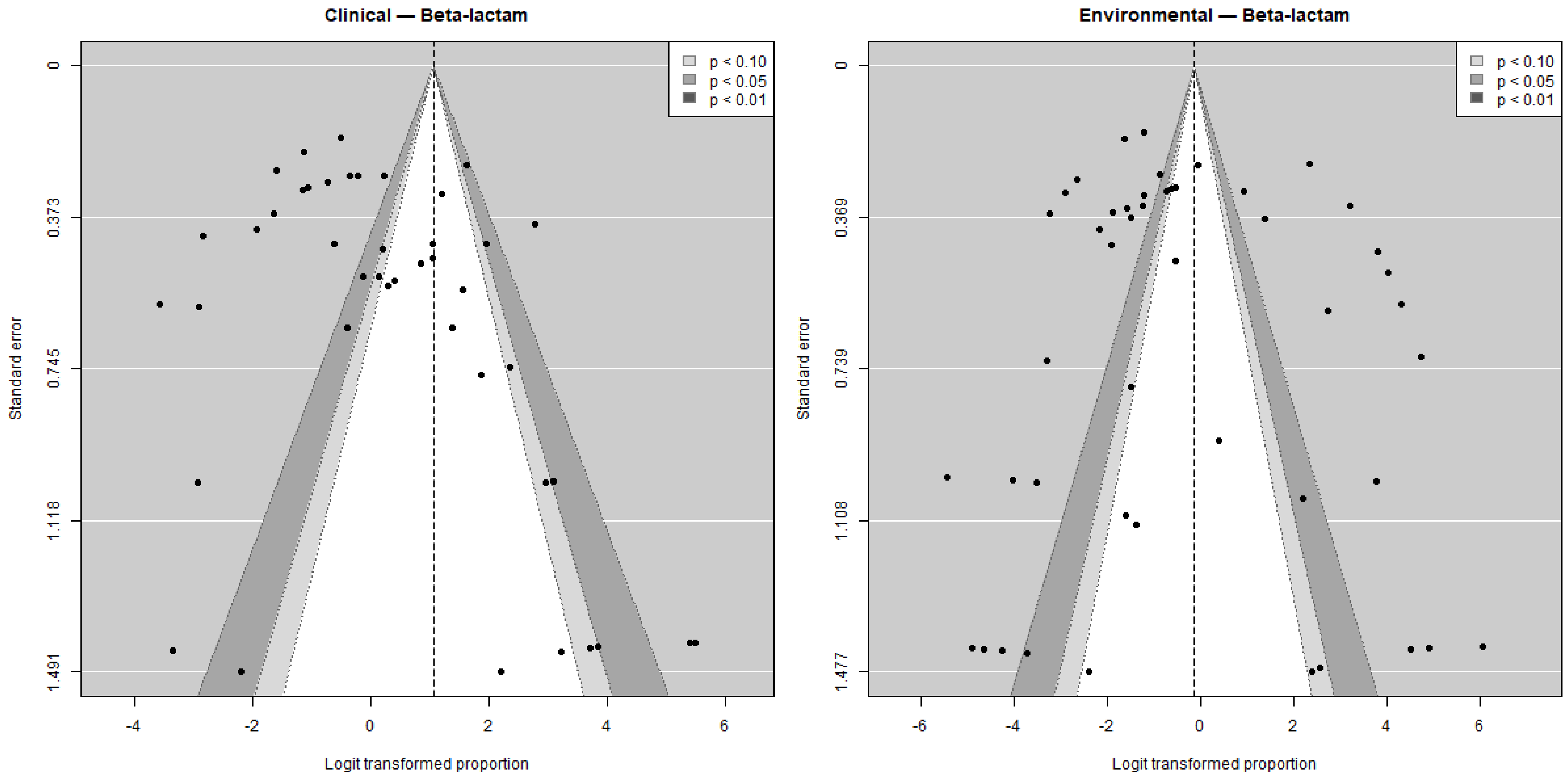

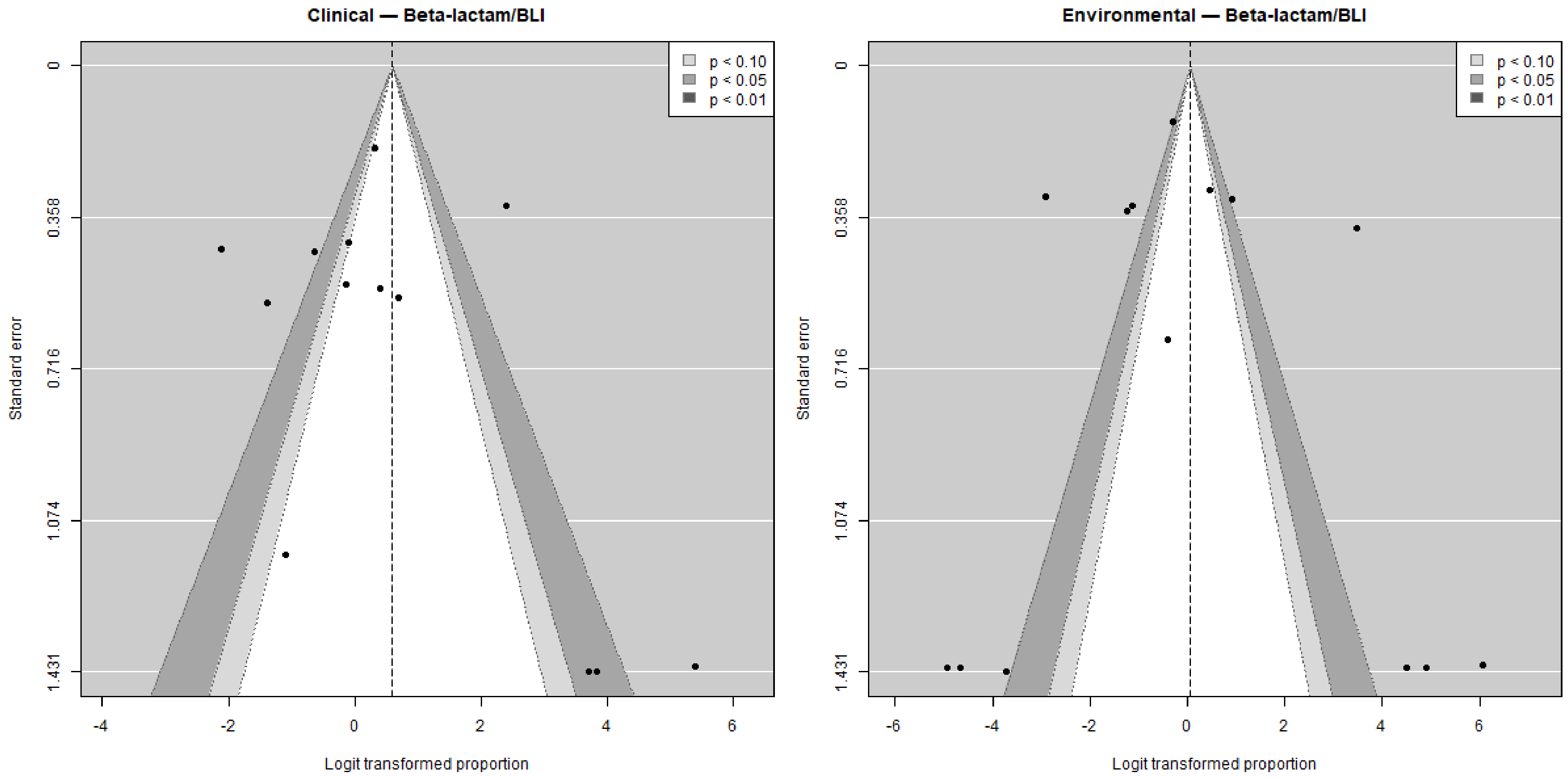

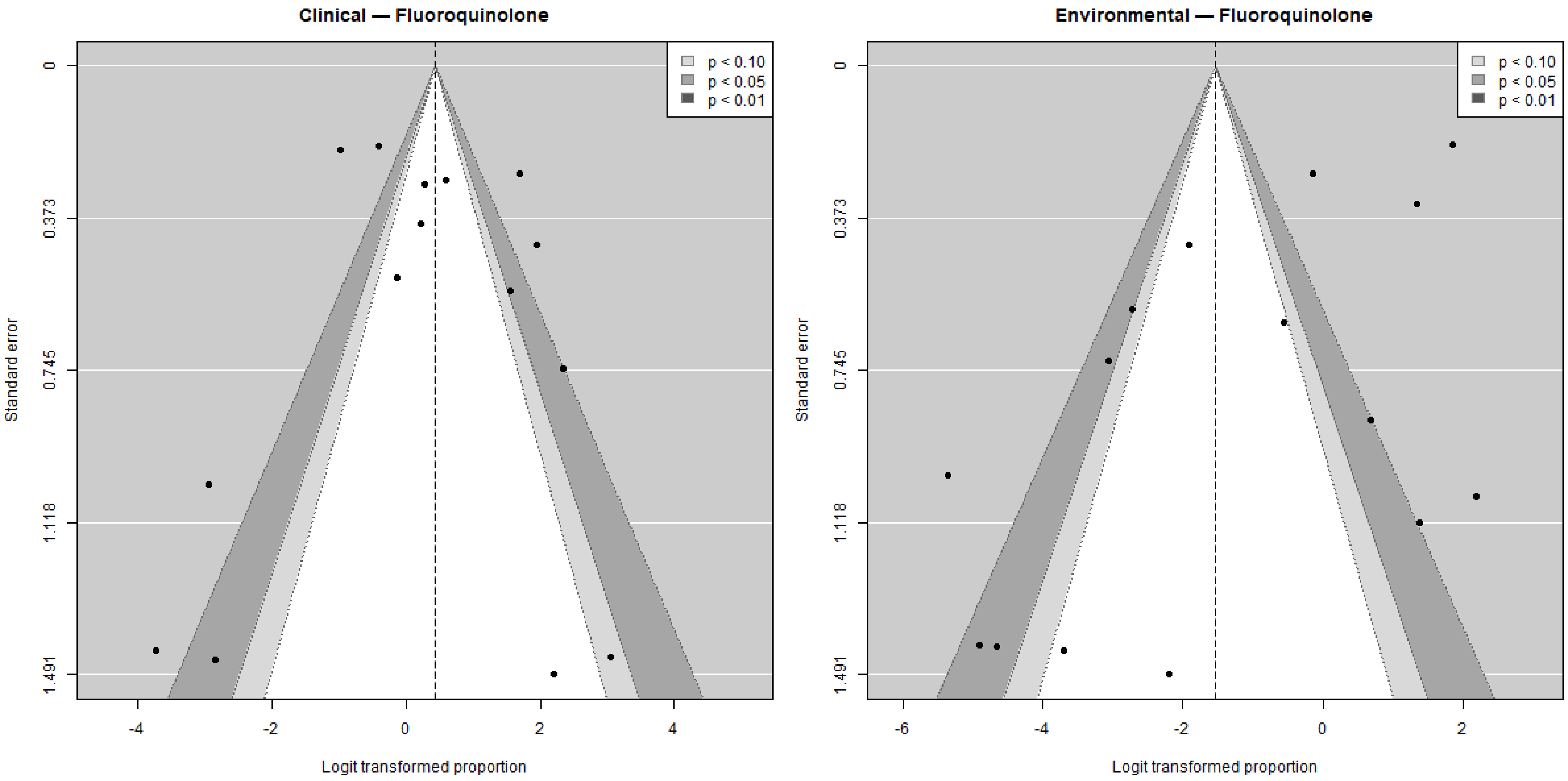

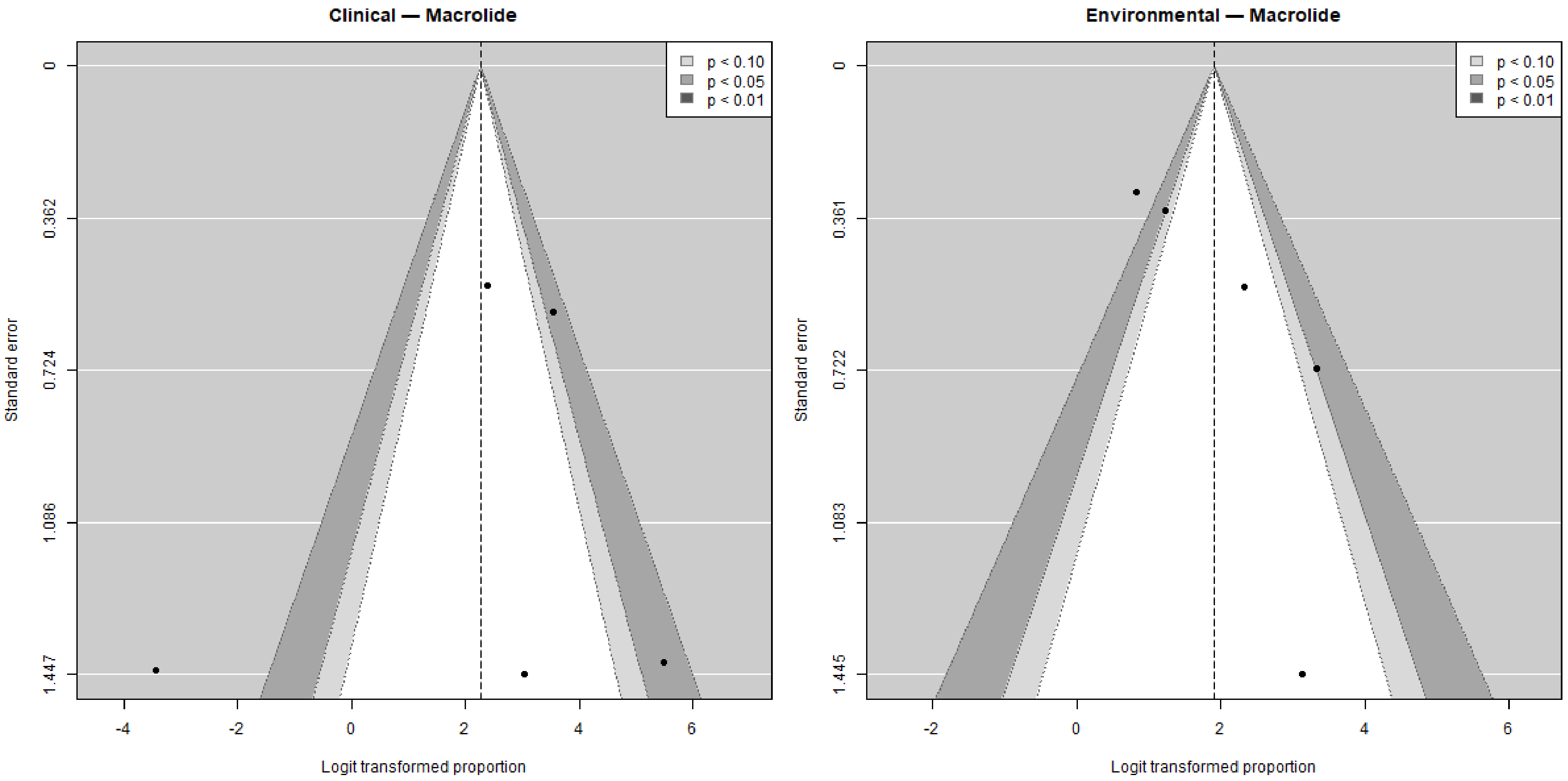

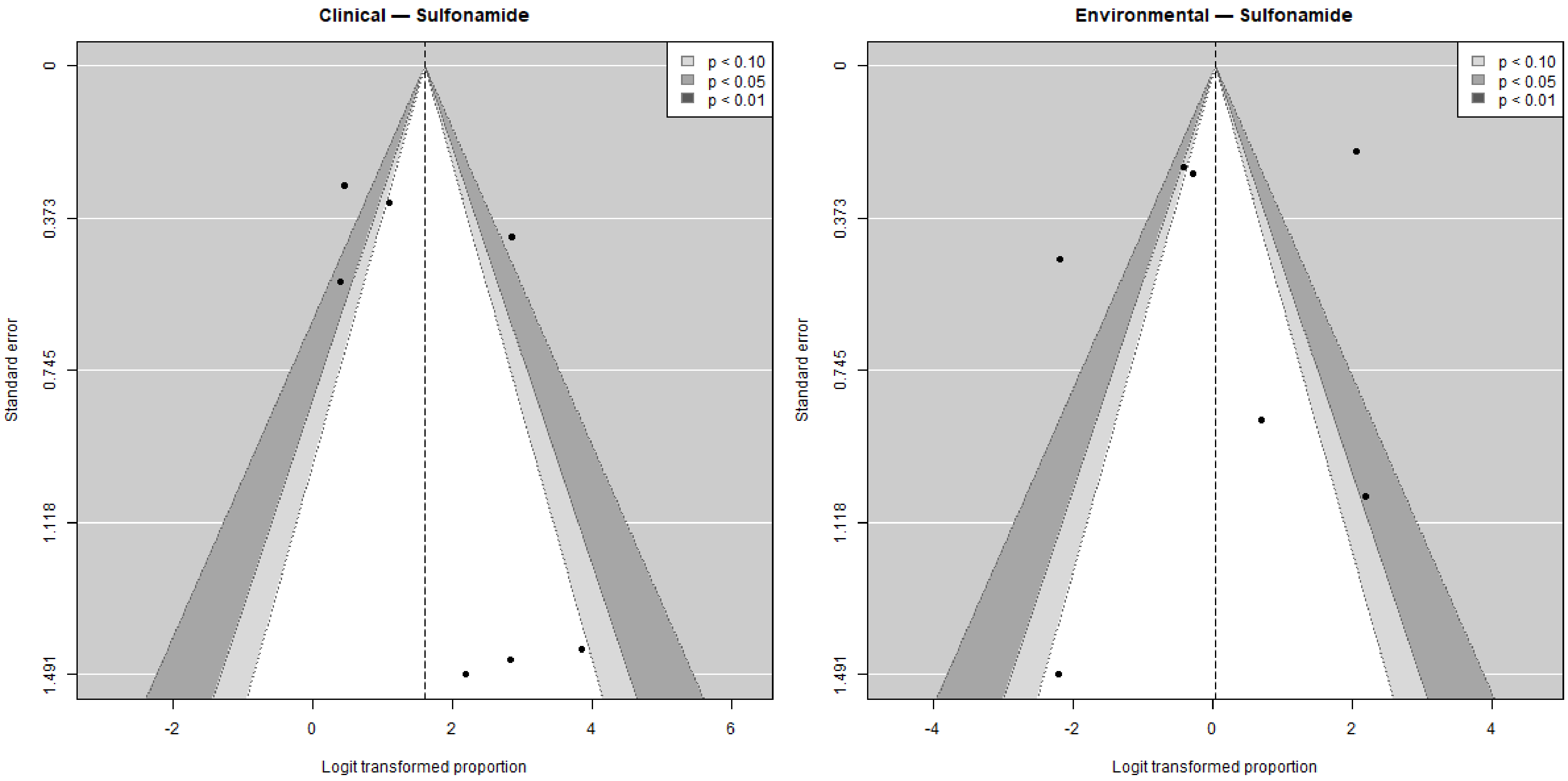

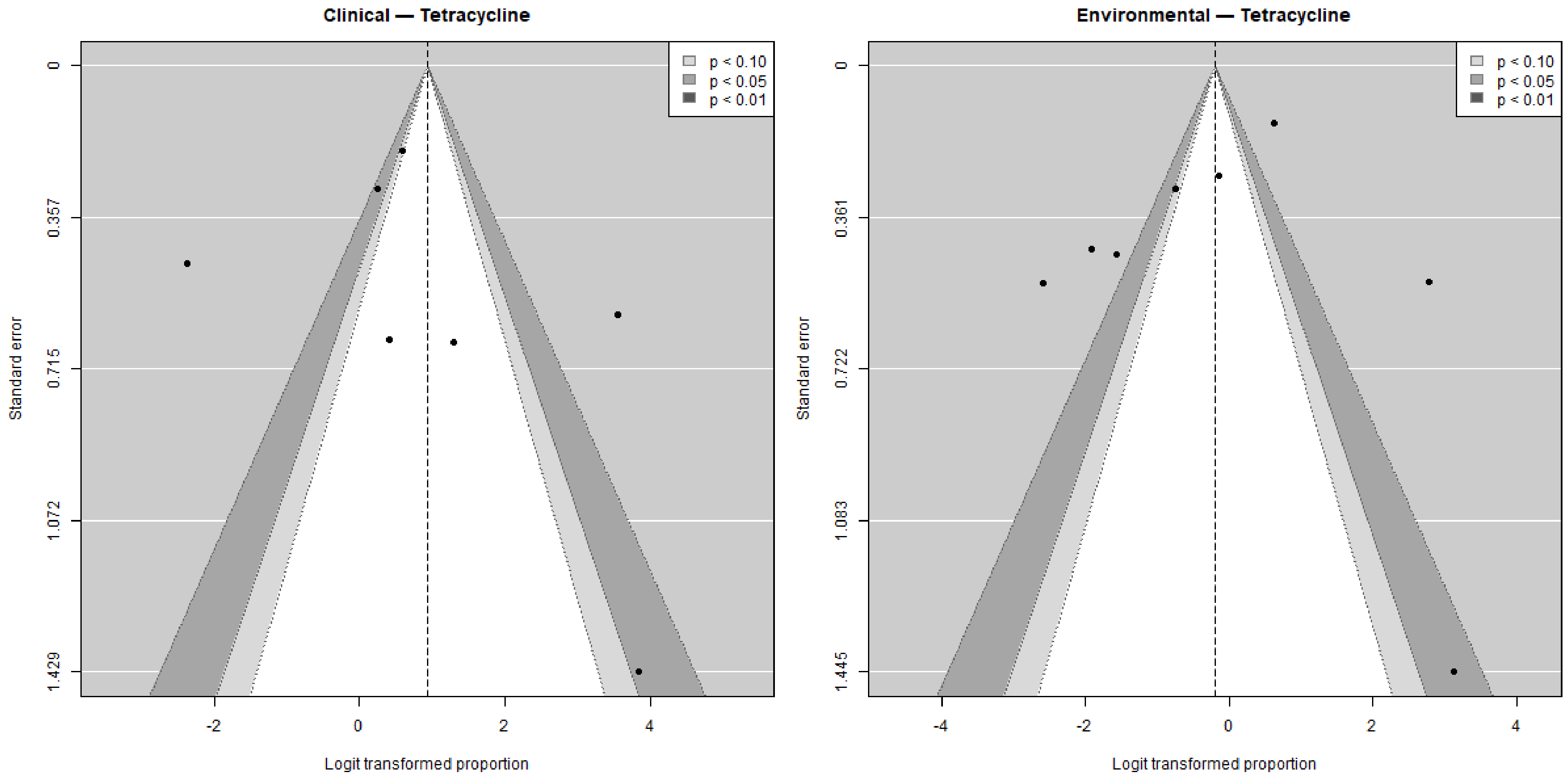


**Supplementary Figure 2.** Funnel plot of the proportion of antibiotic resistance by antibiotic.
