## Supplementary material for "Antimicrobial resistance prevalence in clinical and aquatic environmental ESKAPE: a systematic review with meta-analysis": PRISMA checklist

| **Section and Topic** | **Item #** | **Checklist item** | **Location where item is reported** |
| --- | --- | --- | --- |
| **TITLE** | | |  |
| Title | 1 | Identify the report as a systematic review. | Describe in the title |
| **ABSTRACT** | | |  |
| Abstract | 2 | See the PRISMA 2020 for Abstracts checklist. | It was described in the abstract: Background, objective, data source, methodology, the total number of included studies, the main results, the statistics of meta-analysis with confidence interval, the study limitations, and conclusion.  It was also included the register name and registration number.  It was not included the primary source of funding for the review.  Line numbers: 45-65 |
| **INTRODUCTION** | | |  |
| Rationale | 3 | Describe the rationale for the review in the context of existing knowledge. | Line number: 90 -92: This sentence establishes that AMR is not restricted to clinical settings, justifying interest beyond hospitals.  Line number 95 – 98: This sentence justify why aquatic environments are biologically plausible reservoir of AMR  Line number: 99-101: This explicitly frames the conceptual need for integrating environmental and clinical data  Line number 104 – 107: This justify why ESKAPE pathogens are the focus, not AMR in general  Line number 109-110: This explain the ESKAPE clinical problem and the integration between the two domains.  Line 117 – 119: This is the key gap statement |
| Objectives | 4 | Provide an explicit statement of the objective(s) or question(s) the review addresses. | The objective of this systematic review with meta-analysis was to estimate the prevalence of phenotypic antimicrobial resistance in ESKAPE pathogens isolated from water and wastewater and to compare these estimates with resistance patterns observed in human clinical isolates. |
| **METHODS** | | |  |
| Eligibility criteria | 5 | Specify the inclusion and exclusion criteria for the review and how studies were grouped for the syntheses. | Line number: 146 – 155  **Inclusion criteria**  Original research articles  Reporting phenotypic (culture-based) antimicrobial resistance  In one or more ESKAPE pathogens  With isolates obtained from aquatic environmental matrices (water or wastewater)  And human clinical samples  **Exclusion criteria**  Non-original studies (reviews, editorials, commentaries, protocols)  Case reports without a comparative component  Studies not focused on ESKAPE pathogens  Studies lacking isolates from either environmental or clinical sources  Studies relying exclusively on culture-independent methods without susceptibility testing  Articles not available in full text or lacking a persistent identifier (e.g., DOI) |
| Information sources | 6 | Specify all databases, registers, websites, organisations, reference lists and other sources searched or consulted to identify studies. Specify the date when each source was last searched or consulted. | **Databases searched – Line number 132**  PubMed  Embase  Cochrane Library  **Date last searched – Line number 135**  All databases were searched up to January 14, 2025 |
| Search strategy | 7 | Present the full search strategies for all databases, registers and websites, including any filters and limits used. | The full search strategies for all databased are reported in detail in supplementary table 1. |
| Selection process | 8 | Specify the methods used to decide whether a study met the inclusion criteria of the review, including how many reviewers screened each record and each report retrieved, whether they worked independently, and if applicable, details of automation tools used in the process. | **Line number 135 – 143**  Title and abstracts were screened by four reviewers according to the predefined inclusion and exclusion criteria. The reviewers worked independently and used Zotero for bibliographic management. |
| Data collection process | 9 | Specify the methods used to collect data from reports, including how many reviewers collected data from each report, whether they worked independently, any processes for obtaining or confirming data from study investigators, and if applicable, details of automation tools used in the process. | **Line number 158 – 164 and 169 - 172**  Data were independently extracted by two reviewers and entered into a standardized Microsoft Excel spreadsheet. All data extraction procedures were performed manually, and no automation or software-assisted tools were used in this process. |
| Data items | 10a | List and define all outcomes for which data were sought. Specify whether all results that were compatible with each outcome domain in each study were sought (e.g. for all measures, time points, analyses), and if not, the methods used to decide which results to collect. | For each eligible study, the number of isolates and their corresponding antimicrobial resistance profiles were extracted. The primary outcome was phenotypic antimicrobial resistance prevalence, defined as the proportion of resistant isolates for each specific pathogen–antibiotic combination within each study. **Line number 167 – 172**.  Resistance prevalence was calculated using the total number of isolates tested as the denominator and the number of isolates classified as resistant as the numerator. **Line number 168**  Outcomes were extracted separately for clinical and environmental isolates, allowing direct comparison between sample sources. **Line number 178-179**  Resistance data were collected for each antibiotic in each eligible study. **Line number 167 – 169**  When multiple resistance results were reported within a single study (e.g., multiple antibiotics or pathogens), each unique study–pathogen–antibiotic–source combination was treated as an independent outcome for synthesis. **Line number 171.**  No selection based on time points, statistical significance, or predefined effect direction was applied when collecting outcome data. |
|  | 10b | List and define all other variables for which data were sought (e.g. participant and intervention characteristics, funding sources). Describe any assumptions made about any missing or unclear information. | **Line number 158 - 164**  Bibliographic information, including first author and year of publication.  Pathogen identity, specifying the ESKAPE species investigated.  Geographic location, recorded at the country and continent levels.  Sample source, classified as environmental (water or wastewater) or clinical.  Environmental matrix type, when reported, including effluent-impacted and non-effluent water sources.  Antibiotic tested, recorded at the individual drug level and subsequently grouped by antibiotic class.  No data on participant-level characteristics, interventions, or funding sources of the included studies were systematically extracted, as these were not relevant to the objectives of the review.  When information required for data synthesis was missing, unclear, or ambiguously reported, the study was excluded from quantitative analysis if resistance prevalence could not be reliably derived.  No assumptions or imputations were made to infer missing resistance data, and only explicitly reported phenotypic susceptibility results were included in the synthesis. |
| Study risk of bias assessment | 11 | Specify the methods used to assess risk of bias in the included studies, including details of the tool(s) used, how many reviewers assessed each study and whether they worked independently, and if applicable, details of automation tools used in the process. | No formal risk of bias tool was applied to the included studies.  This decision was based on the observational.  Given that the primary outcome was phenotypic antimicrobial resistance prevalence derived from aggregated isolate-level data, and not effect estimates from comparative or interventional designs, standard risk-of-bias tools (e.g., for randomized or non-randomized clinical studies) were not considered appropriate  Source of methodological variability and potential bias were explored indirectly through subgroup analysis, heterogeneity statistics (τ², I²), and sensitivity considerations during data synthesis. **Line number: 173 - 179** |
| Effect measures | 12 | Specify for each outcome the effect measure(s) (e.g. risk ratio, mean difference) used in the synthesis or presentation of results. | For the primary outcome, phenotypic antimicrobial resistance prevalence, the effect measure used was the proportion of resistant isolates for each pathogen–antibiotic–source combination. **Line number: 167 - 172**  Resistance prevalence was expressed as a pooled proportion with corresponding 95% confidence intervals (CIs). **Line number 173 - 176**  For meta-analytic synthesis, proportions were logit-transformed and analyzed using binomial–normal generalized linear mixed models (GLMMs) to account for within- and between-study variability. **Line number: 173 – 175.**  Summary estimates were back-transformed to the original proportion scale for presentation of results. **Line number: 175 – 176.**  Comparative presentation between clinical and environmental sources was based on stratified pooled prevalence estimates, rather than relative effect measures**. Line number: 178-179**  Between-study heterogeneity was quantified using τ², I², and H² statistics and reported alongside pooled estimates. **Line number: 176-177** |
| Synthesis methods | 13a | Describe the processes used to decide which studies were eligible for each synthesis (e.g. tabulating the study intervention characteristics and comparing against the planned groups for each synthesis (item #5)). | Only studies reporting sufficient data on the number of isolates tested and their corresponding antimicrobial resistance profiles were included. Data were tabulated by pathogen and by the absolute number of isolates exhibiting resistance to each specific antimicrobial agent. **Line number: 162 - 164** |
|  | 13b | Describe any methods required to prepare the data for presentation or synthesis, such as handling of missing summary statistics, or data conversions. | Resistance data were standardized by expressing outcomes as proportions of resistant isolates. **Line number: 167 – 169.**  When resistance results were reported for multiple antibiotics or pathogens within the same study, each unique study–pathogen–antibiotic–source combination was treated as an independent data point for synthesis. **Line number 171 – 172.**  Proportion data were logit-transformed before meta-analysis to stabilize variances and accommodate the bounded nature of prevalence estimates, with pooled results subsequently back-transformed to the original proportion scale for presentation. **Line number: 175 - 176**  Except for one study, all included studies reported complete information on the number of isolates tested and the number of resistant isolates. The study lacking complete isolate counts was excluded from the quantitative synthesis but retained for qualitative description. **Line number: 193 – 195**.  Antibiotics were grouped into antibiotic classes for subgroup analyses, and results were stratified by clinical and environmental sources. **Line number: 178 - 179** |
|  | 13c | Describe any methods used to tabulate or visually display results of individual studies and syntheses. | Characteristics and extracted data from individual studies were tabulated in summary tables, including pathogen, sample source (clinical or environmental), antibiotic tested, number of isolates evaluated, and number of resistant isolates**. Supplementary table 3.**  Results of quantitative syntheses were visually displayed using forest plots, presenting pooled resistance prevalence estimates with corresponding 95% confidence intervals for each antibiotic and antibiotic class. Figure 3 and **Supplementary figure 2.**  Forest plots were stratified by sample source (clinical versus environmental) to facilitate direct comparison between ecological compartments. **Supplementary figure 3.**  Antibiotics within each forest plot were ordered by overall mean resistance prevalence to enhance interpretability. **Figure 3**  Contour-enhanced funnel plots of logit-transformed proportions were used to visually assess small-study effects when at least five independent studies were available for a given antibiotic class. **Supplementary figure 3.**  All tables and figures were generated using R (version 4.2.2), and graphical outputs were produced using standard functions from the metafor and meta packages. |
|  | 13d | Describe any methods used to synthesize results and provide a rationale for the choice(s). If meta-analysis was performed, describe the model(s), method(s) to identify the presence and extent of statistical heterogeneity, and software package(s) used. | Pooled resistance prevalence was estimated using binomial–normal generalized linear mixed models (GLMMs) with a logit link function, treating each unique study–pathogen–antibiotic–source combination as an independent effect size. This random-effects framework was chosen a priori to account for both within-study binomial variability and between-study heterogeneity, which were expected given the diversity of study designs, geographic locations, sample matrices (clinical vs. environmental), and antimicrobial agents assessed.  Pooled logit-transformed estimates and their 95% confidence intervals were back-transformed to the proportion scale for interpretation and presentation of resistance prevalence.  To identify the presence and extent of statistical heterogeneity, we applied Cochran’s Q test, estimated the between-study variance (τ²) using maximum likelihood, and quantified heterogeneity using the I² and H² statistics. Tests of homogeneity (τ² = 0) were performed using Wald and likelihood-ratio tests.  All quantitative syntheses and heterogeneity analyses were carried out in R (version 4.2.2) using the metafor and meta packages.  **Lines 173–185** |
|  | 13e | Describe any methods used to explore possible causes of heterogeneity among study results (e.g. subgroup analysis, meta-regression). | Potential sources of heterogeneity were explored through a priori subgroup analyses, conducted when a sufficient number of studies were available per category. Subgroups were defined according to data source (environmental versus clinical isolates), bacterial species, antimicrobial class, and geographic region, reflecting key epidemiological and methodological differences across the included studies.  Subgroup-specific pooled resistance estimates were calculated using the same random-effects GLMM framework applied in the main meta-analyses to ensure methodological consistency. Differences between subgroups were assessed by comparing pooled estimates and their confidence intervals, as well as changes in heterogeneity metrics (τ² and I²) across strata.  Formal meta-regression analyses were not performed due to the limited number of studies available for several pathogen–antibiotic combinations and the high variability in study-level covariates, which could compromise model stability and interpretability.  **Line number: 173 - 185** |
|  | 13f | Describe any sensitivity analyses conducted to assess robustness of the synthesized results. | Robustness was assessed qualitatively by checking whether the main resistance patterns remained consistent across stratified analyses (clinical vs. environmental, antibiotic class, individual antibiotics, and water matrices). **Line number: 173 - 185** |
| Reporting bias assessment | 14 | Describe any methods used to assess risk of bias due to missing results in a synthesis (arising from reporting biases). | Risk of bias due to missing results (small-study effects) was assessed using contour-enhanced funnel plots and Egger’s regression test for antibiotic classes with ≥ 5 studies, stratified by clinical and environmental samples. Significant asymmetry was detected for some classes, but as discussed in the manuscript, these patterns were interpreted mainly as reflecting methodological heterogeneity and uneven antibiotic representation, rather than classic publication bias.  **Line number: 181 - 184** |
| Certainty assessment | 15 | Describe any methods used to assess certainty (or confidence) in the body of evidence for an outcome. | Certainty in the body of evidence was assessed qualitatively through evaluation of consistency of findings across stratified analyses, magnitude and direction of pooled estimates, degree of between-study heterogeneity, and methodological limitations discussed in the manuscript. These considerations supported confidence in the overall patterns observed while warranting cautious interpretation of precise prevalence estimates.  **Line number: 218 – 222/ 329 – 333/ 335 – 345 / 384 - 403** |
| **RESULTS** | | |  |
| Study selection | 16a | Describe the results of the search and selection process, from the number of records identified in the search to the number of studies included in the review, ideally using a flow diagram. | A total of 304 records were identified through database searching. After screening titles, abstracts, and full texts, 18 studies met the eligibility criteria and were included in the review and meta-analysis. The study selection process and reasons for exclusion are presented in the PRISMA flow diagram. **Line number 189 – 195 and Figure 1.** |
|  | 16b | Cite studies that might appear to meet the inclusion criteria, but which were excluded, and explain why they were excluded. | Several studies that initially appeared to meet the inclusion criteria were excluded after full-text review. The main reasons for exclusion were the absence of phenotypic antimicrobial susceptibility data, exclusive use of genomic or metagenomic approaches without culture-based resistance results, pre-selection of resistant isolates that prevented prevalence estimation, or study populations outside the predefined scope (e.g., ICU-only or non-comparative designs). **Line number: 189 – 195 and Figure 1.**  One study was considered qualitatively relevant but was excluded from the quantitative synthesis due to the lack of isolate incidence data required to calculate resistance prevalence. Detailed reasons for exclusion are summarized in the PRISMA flow diagram. **Line number: 189 – 195 and Figure 1.** |
| Study characteristics | 17 | Cite each included study and present its characteristics. | A total of 18 studies were included in the systematic review and meta-analysis. The characteristics of each included study—comprising first author and year of publication, country and continent, ESKAPE pathogen(s) investigated, sample origin (clinical and/or environmental), water matrix when applicable, number of isolates analyzed, and antibiotics tested—are summarized in **Supplementary Tables 2 and 3.** |
| Risk of bias in studies | 18 | Present assessments of risk of bias for each included study. | Potential sources of bias were assessed qualitatively based on study design, sampling strategies, isolate selection, laboratory methods, and completeness of antimicrobial susceptibility reporting. Methodological heterogeneity related to sample matrices, selective isolation procedures, and uneven antibiotic testing was explicitly considered during data synthesis and interpretation of results. These aspects are described in the study selection criteria and exclusion rationale (**Line number: 189 - 195**), detailed in the characterization of included studies **(Line number 218 – 227)**. |
| Results of individual studies | 19 | For all outcomes, present, for each study: (a) summary statistics for each group (where appropriate) and (b) an effect estimate and its precision (e.g. confidence/credible interval), ideally using structured tables or plots. | For all outcomes, study-level summary statistics were extracted as the number of resistant isolates and total isolates tested for each pathogen–antibiotic–source combination. These data were used to calculate study-specific resistance proportions. **Supplementary table 2.**  Individual study estimates and their corresponding 95% confidence intervals were incorporated into the quantitative synthesis and are presented visually in forest plots, stratified by antibiotic class, individual antibiotics, and sample origin (clinical vs environmental). **Supplementary figure 2.** |
| Results of syntheses | 20a | For each synthesis, briefly summarise the characteristics and risk of bias among contributing studies. | Included studies were observational and laboratory-based, reporting phenotypic antimicrobial resistance in ESKAPE pathogens. Studies differed substantially in sample size, geographic setting, environmental matrices, antibiotic panels, and laboratory methods. Risk of bias was assessed qualitatively and was mainly related to sampling strategies, isolate pre-selection (e.g., selective media), heterogeneity of laboratory procedures, and incomplete susceptibility reporting, contributing to the high between-study heterogeneity observed. **Line number: 199-205/ 218 – 227/ 276-281.** |
|  | 20b | Present results of all statistical syntheses conducted. If meta-analysis was done, present for each the summary estimate and its precision (e.g. confidence/credible interval) and measures of statistical heterogeneity. If comparing groups, describe the direction of the effect. | All statistical syntheses showed high heterogeneity. The overall pooled resistance prevalence was 0.46 (95% CI 0.36–0.57; I² = 98.8%). Resistance was higher in clinical isolates (0.67, 95% CI 0.55–0.77) than in environmental isolates (0.24, 95% CI 0.14–0.39), indicating a consistent direction toward higher resistance in clinical settings. Subgroup analyses by antibiotic class and water matrix showed similar patterns, with higher resistance in effluent-impacted waters, while heterogeneity remained substantial. **Line number: 218-227.**  The direction of effect consistently indicated higher resistance in clinical settings. **Line number 223-227** |
|  | 20c | Present results of all investigations of possible causes of heterogeneity among study results. | Potential sources of heterogeneity were investigated through stratified analyses by sample origin (clinical vs environmental), antibiotic class, individual antibiotics, and environmental water matrices (effluent-impacted vs non-effluent). Heterogeneity remained high across all analyses (I² ≈ 98%), indicating substantial true variability between studies. Variability was primarily attributed to differences in sampling matrices, laboratory isolation methods (including use of selective media), and uneven representation of antibiotics across studies, rather than random error. These factors were identified as the main contributors to between-study heterogeneity. **Line number: 218-227/ 262-273/ 276-281.** |
|  | 20d | Present results of all sensitivity analyses conducted to assess the robustness of the synthesized results. | Robustness of the synthesized results was instead assessed through multiple stratified analyses (by sample origin, antibiotic class, individual antibiotics, and water matrices) and by examining the consistency and direction of pooled estimates across these analyses. The persistence of overall patterns despite substantial heterogeneity supports the robustness of the main findings, while warranting cautious interpretation of precise prevalence estimates. **Line number 230-273.** |
| Reporting biases | 21 | Present assessments of risk of bias due to missing results (arising from reporting biases) for each synthesis assessed. | Risk of bias due to missing results was assessed through visual inspection of funnel plots and Egger’s regression test for syntheses including at least five studies. Evidence of funnel plot asymmetry was observed for selected antibiotic classes; however, these patterns were interpreted as reflecting methodological heterogeneity and uneven representation of individual antibiotics within classes, rather than selective non-reporting of studies. No clear evidence of systematic reporting bias affecting the overall direction of results was identified across syntheses. **Line number 276 – 281/ Supplementary figure 3/ Supplementary table 4 and 5.** |
| Certainty of evidence | 22 | Present assessments of certainty (or confidence) in the body of evidence for each outcome assessed. | For each outcome, certainty in the body of evidence was assessed qualitatively, based on the consistency and direction of pooled prevalence estimates across stratified analyses, the magnitude of between-study heterogeneity, and methodological limitations of the included studies. Across outcomes, results showed consistent directional patterns (e.g., higher resistance in clinical compared with environmental isolates), supporting confidence in the overall trends, while substantial heterogeneity and variability in study methods warranted cautious interpretation of precise prevalence estimates. **Line number 276-281** |
| **DISCUSSION** | | |  |
| -Discussion | 23a | Provide a general interpretation of the results in the context of other evidence. | Overall, antimicrobial resistance in ESKAPE pathogens was more prevalent in clinical than in aquatic environmental isolates, consistent with previous evidence. Aquatic environments, particularly effluent-impacted waters, appear to function as important reservoirs of resistance, supporting a One Health perspective despite substantial heterogeneity across studies. **Line number 327-403** |
|  | 23b | Discuss any limitations of the evidence included in the review. | The evidence is limited by substantial methodological and ecological heterogeneity, variability in sampling and laboratory methods, selective isolation approaches, and uneven reporting of antimicrobial susceptibility data, which restricts comparability and precision of pooled estimates. **Line number 329-356.** |
|  | 23c | Discuss any limitations of the review processes used. | The review was limited to published, culture-based studies and was constrained by heterogeneity in study design and reporting, which restricted standardization and some comparisons. **Line number: 329-333** |
|  | 23d | Discuss implications of the results for practice, policy, and future research. | The results support strengthened antimicrobial stewardship in clinical settings and inclusion of environmental surveillance in AMR policies. Future research should prioritize standardized methodologies to improve comparability and reduce heterogeneity. **Line number 329-403** |
| **OTHER INFORMATION** | | |  |
| Registration and protocol | 24a | Provide registration information for the review, including register name and registration number, or state that the review was not registered. | https://www.crd.york.ac.uk/PROSPERO/view/CRD420251020930, CRD420251020930. **Line number: 125-127** |
|  | 24b | Indicate where the review protocol can be accessed, or state that a protocol was not prepared. | The review protocol was prospectively registered in PROSPERO and is publicly available at:  https://www.crd.york.ac.uk/PROSPERO/view/CRD420251020930  (Registration number: CRD420251020930). |
|  | 24c | Describe and explain any amendments to information provided at registration or in the protocol. | The protocol was refined after registration. Although initially focused on resistance genes, most included studies did not evaluate gene-level resistance. The review therefore focused on phenotypic antimicrobial resistance, without changes to eligibility criteria, searches, or study selection. |
| Support | 25 | Describe sources of financial or non-financial support for the review, and the role of the funders or sponsors in the review. | This study was funded through reparation funds originating from the Vale mining company. The funding source had no involvement in the design of the review, data collection, analysis, interpretation of findings, or writing of the manuscript. |
| Competing interests | 26 | Declare any competing interests of review authors. | The authors declare that they have no known competing financial or non-financial interests that could have influenced the work reported in this paper. |
| Availability of data, code and other materials | 27 | Report which of the following are publicly available and where they can be found: template data collection forms; data extracted from included studies; data used for all analyses; analytic code; any other materials used in the review. | Data extracted from the included studies and used in all analyses are publicly available in **Supplementary Table 2**. No separate public repository was used for the analytic code or data extraction templates. |

*From:*  Page MJ, McKenzie JE, Bossuyt PM, Boutron I, Hoffmann TC, Mulrow CD, et al. The PRISMA 2020 statement: an updated guideline for reporting systematic reviews. BMJ 2021;372:n71. doi: 10.1136/bmj.n71. This work is licensed under CC BY 4.0. To view a copy of this license, visit <https://creativecommons.org/licenses/by/4.0/>
